# Association of Structural Forms of 17q21.31 with the Risk of Progressive Supranuclear Palsy and *MAPT* Sub-haplotypes

**DOI:** 10.1101/2024.02.26.24303379

**Authors:** Hui Wang, Timothy S Chang, Beth A Dombroski, Po-Liang Cheng, Ya-Qin Si, Albert Tucci, Vishakha Patil, Leopoldo Valiente-Banuet, Kurt Farrell, Catriona Mclean, Laura Molina-Porcel, Rajput Alex, Peter Paul De Deyn, Nathalie Le Bastard, Marla Gearing, Laura Donker Kaat, John C Van Swieten, Elise Dopper, Bernardino F Ghetti, Kathy L Newell, Claire Troakes, Justo G de Yébenes, Alberto Rábano-Gutierrez, Tina Meller, Wolfgang H Oertel, Gesine Respondek, Maria Stamelou, Thomas Arzberger, Sigrun Roeber, Ulrich Müller, Franziska Hopfner, Pau Pastor, Alexis Brice, Alexandra Durr, Isabelle Le Ber, Thomas G Beach, Geidy E Serrano, Lili-Naz Hazrati, Irene Litvan, Rosa Rademakers, Owen A Ross, Douglas Galasko, Adam L Boxer, Bruce L Miller, Willian W Seeley, Vivianna M Van Deerlin, Edward B Lee, Charles L White, Huw R Morris, Rohan de Silva, John F Crary, Alison M Goate, Jeffrey S Friedman, Yuk Yee Leung, Giovanni Coppola, Adam C Naj, Li-San Wang, PSP genetics study group, Dennis W Dickson, Günter U Höglinger, Jung-Ying Tzeng, Daniel H Geschwind, Gerard D Schellenberg, Wan-Ping Lee

## Abstract

**Importance:** The chromosome 17q21.31 region, containing a 900 Kb inversion that defines H1 and H2 haplotypes, represents the strongest genetic risk locus in progressive supranuclear palsy (PSP). In addition to H1 and H2, various structural forms of 17q21.31, characterized by the copy number of α, β, and γ duplications, have been identified. However, the specific effect of each structural form on the risk of PSP has never been evaluated in a large cohort study.

**Objective:** To assess the association of different structural forms of 17q.21.31, defined by the copy numbers of α, β, and γ duplications, with the risk of PSP and *MAPT* sub-haplotypes.

**Design, setting, and participants:** Utilizing whole genome sequencing data of 1,684 (1,386 autopsy confirmed) individuals with PSP and 2,392 control subjects, a case-control study was conducted to investigate the association of copy numbers of α, β, and γ duplications and structural forms of 17q21.31 with the risk of PSP. All study subjects were selected from the Alzheimer’s Disease Sequencing Project (ADSP) Umbrella NG00067.v7. Data were analyzed between March 2022 and November 2023.

**Main outcomes and measures:** The main outcomes were the risk (odds ratios [ORs]) for PSP with 95% CIs. Risks for PSP were evaluated by logistic regression models.

**Results:** The copy numbers of α and β were associated with the risk of PSP only due to their correlation with H1 and H2, while the copy number of γ was independently associated with the increased risk of PSP. Each additional duplication of γ was associated with 1.10 (95% CI, 1.04-1.17; *P* = 0.0018) fold of increased risk of PSP when conditioning H1 and H2. For the H1 haplotype, addition γ duplications displayed a higher odds ratio for PSP: the odds ratio increases from 1.21 (95%CI 1.10-1.33, *P* = 5.47 × 10^-5^) for H1β1γ1 to 1.29 (95%CI 1.16-1.43, *P* = 1.35 × 10^-6^) for H1β1γ2, 1.45 (95%CI 1.27-1.65, *P* = 3.94 × 10^-8^) for H1β1γ3, and 1.57 (95%CI 1.10-2.26, *P* = 1.35 × 10^-2^) for H1β1γ4. Moreover, H1β1γ3 is in linkage disequilibrium with H1c (R^2^ = 0.31), a widely recognized *MAPT* sub-haplotype associated with increased risk of PSP. The proportion of *MAPT* sub-haplotypes associated with increased risk of PSP (i.e., H1c, H1d, H1g, H1o, and H1h) increased from 34% in H1β1γ1 to 77% in H1β1γ4.

**Conclusions and relevance:** This study revealed that the copy number of γ was associated with the risk of PSP independently from H1 and H2. The H1 haplotype with more γ duplications showed a higher odds ratio for PSP and were associated with *MAPT* sub-haplotypes with increased risk of PSP. These findings expand our understanding of how the complex structure at 17q21.31 affect the risk of PSP.

**Key Points:** *Question:* Do large copy number variations (i.e., α, β, and γ) inside 17q21.31 contribute to the risk of progressive supranuclear palsy (PSP) independently from the H1 and H2 haplotypes? Do structural forms of 17q21.31, characterized by combinations of α, β, and γ, present divergent risk to the development of PSP? Are structural forms of 17q21.31 associated with *MAPT* sub-haplotypes, such as H1c?

*Findings:* In this case-control study of 1,684 individuals with PSP and 2,392 control subjects, the copy number of γ duplication was independently associated with the risk of the disease. H1 haplotypes with more γ duplications (H1β1γ2, H1β1γ3, and H1β1γ4) displayed a higher odds ratio for PSP when compared to H1β1γ1. Notably, H1β1γ3 was observed to be in linkage disequilibrium with H1c, a widely recognized *MAPT* sub-haplotype associated with PSP.

*Meaning:* The association between the H1 and H2 haplotypes and PSP involves multiple contributing factors, including the copy number of γ duplication.

## Introduction

Progressive supranuclear palsy (PSP) is a neurodegenerative disease with two characteristic clinical features, i.e. postural instability and ocular motor abnormalities^1^. Other symptoms and signs, such as cognitive disfunction and problems with swallowing, vary and can get worse over time depending on the distribution of pathology and severity of diseases^2,3^. The main pathology of PSP is the accumulation of tau in the brain, leading to the presence of neurofibrillary tangles and threads along with tufted astrocytes and oligodendroglial coiled bodies^4,5^. An isoform of tau harboring 4 repeats (4R) of microtubule-binding domain is particularly prominent in these tau aggregates^6^.

The most recognized genetic risk locus for PSP is situated on 17q21.31. The region can be divided into two major haplotypes, H1 and H2, characterized by a 900 Kb inversion found in 20% of Europeans^7^. Individuals carrying the H1 haplotype, compared to the H2 haplotype, are more likely to develop PSP, with an estimated odds ratio (OR) around 5 in Europeans^8,9^. The association was identified through various variants in linkage disequilibrium (LD) with H1 and H2, such as a dinucleotide repeat (TG)_n_ in *MAPT* intron 9^10^, a 238-bp deletion in *MAPT* intron 9^7^, and a multitude of single nucleotide variants (SNVs) in this region^8,9^. However, the causal variants underlying the association remain unclear due to numerous SNVs, short insertion/deletions (indels), and structural variations (SVs) introduced by complex genomic rearrangements in 17q.21.31.

Based on LD structure in the *MAPT* region, H1 and H2 haplotypes can be further categorized into more than 20 common sub-haplotypes^11–13^. Among them, H1c, H1d, H1g, and H1o were significantly associated with the increased risk of PSP^11,13^. Particularly, H1c and H1d are nominally associated with measures of severity of tau pathology in PSP cases^11^. However, these sub-haplotypes were inferred from LD structure limited to *MAPT* gene (∼150 Kb) and do not represent the intricacies of structural forms on 17q21.31 (∼1.5 Mb). To address this limitation, we constructed the ten structural forms of 17q21.31, characterized by the copy numbers of α, β, and γ (150Kb, 300Kb, and 218Kb; eFigure 1 in **Supplement**)^14,15^, and evaluated their association with the risk of PSP in this study. We found that the H1 haplotype with more copies γ duplication showed an increased risk of PSP. The result is in accordance with the higher portion of H1c and other *MAPT* sub-haplotypes associated with increased risk of PSP in the H1 haplotypes with more copies of γ. Through this study, we gained a better understanding how the haplotypic structure of 17q.21.31 correlates with the risk of PSP.

## Methods

### Study subjects and quality control

All study subjects and WGS data are available on The National Institute on Aging Genetics of Alzheimer’s Disease Data Storage Site (NIAGADS)^16^ under Alzheimer’s Disease Sequencing Project (ADSP) Umbrella NG00067.v7^17^. We inferred ancestry of subjects by GRAF-pop^18^ and selected 4,618 subjects (1,797 cases and 2,821 controls) of European ancestry for analysis. WGS were performed at 30x coverage (e**Table 1**).

Among 4,618 samples, we filtered 183 samples with abnormally low reads mapped (aligned read depth < 1.7x) to α, β or γ region (eFigure 2 in **Supplement**) and ten samples with high genotyping missing rate (> 0.05). Next, 244 related samples inferred by KING^19^ (duplicates, monozygotic twins, parent-offsprings, full-siblings, and 2^nd^ degree relatives) were removed.

We used the 238-bp deletion between exons 9 and 10 of *MAPT*^7^ to determine the H1 and H2 haplotypes of each sample. The genotype calls of the 238-bp deletion were obtained from our previous SV work^20^. 75 subjects were removed due to missing or failed genotype of the 238-bp deletion. Given the specification of H1/H2 genotype, determined by the 238 bp deletion, and the copy numbers of α, β and γ, we can ascertain the ten structural forms (eFigure 1 in **Supplement**) in each individual. We removed 30 individuals (eFigure 3 and eFigure 4 in **Supplement**) since their structural forms could not be decided based on the copy numbers of α, β and γ. This discordance might be due to subjects carrying undiscovered structural forms or genotyping errors on the copy numbers of α, β and γ. As a result, 4,076 subjects (**Table 1**; N_PSP_ = 1,684, N_control_ = 2,392) remained for statistical analyses in this study. Of the 1,684 individuals with PSP, 1,386 were autopsy confirmed.

**Table 1.** Characteristics of PSP cases and controls.

|  | **Overall**<br>**(N = 4,076)** | **PSP**<br>**(N = 1,684)** | **Control**<br>**(N = 2,392)** |
| --- | --- | --- | --- |
| **Age<sup>a</sup> (SD)** | 78.49 (8.50) | 68.03 (8.17) | 81.04 (6.37) |
| **Sex (%)** |  |  |  |
| Female | 2168 (53.19%) | 739 (43.88%) | 1429 (59.74%) |
| Male | 1908 (46.81%) | 945 (56.12%) | 963 (40.26%) |
| **H1/H2 status<sup>b</sup> (%)** |  |  |  |
| H1H1 | 2958 (72.57%) | 1511 (89.73%) | 1447 (60.49%) |
| H1H2 | 975 (23.92%) | 168 (9.98%) | 807 (33.74%) |
| H2H2 | 143 (3.51%) | 5 (0.30%) | 138 (5.77%) |
| **Structural forms of 17q21.31<sup>c</sup> (%)** |  |  |  |
| H1β1γ1 | 2446 (30.00%) | 1097 (32.57%) | 1349 (28.20%) |
| H1β1γ2 | 1552 (19.04%) | 739 (21.94%) | 813 (16.99%) |
| H1β1γ3 | 987 (12.11%) | 496 (14.73%) | 491 (10.26%) |
| H1β1γ4 | 126 (1.55%) | 65 (1.93%) | 61 (1.28%) |
| H1β2γ1 | 1716 (21.05%) | 774 (22.98%) | 942 (19.69%) |
| H1β3γ1 | 64 (0.79%) | 19 (0.56%) | 45 (0.94%) |
| H2α1γ1 | 7 (0.09%) | 1 (0.03%) | 6 (0.13%) |
| H2α1γ2 | 99 (1.21%) | 14 (0.42%) | 85 (1.78%) |
| H2α2γ1 | 33 (0.40%) | 2 (0.06%) | 31 (0.65%) |
| H2α2γ2 | 1122 (13.76%) | 161 (4.78%) | 961 (20.09%) |

<sup>a</sup>1,130 PSP cases and 111 controls have missing age.
<sup>b</sup>H1/H2 status was determined by the genotype of a 238-bp H2 tagging deletion<sup>7</sup>.
<sup>c</sup>Structural forms of 17q21.31 were inferred by expectation-maximization (EM) algorithm.

### Calling the copy numbers α, β and γ duplications from WGS

The genomic coordinates on HG38 of α (chr17:46135415-46289349), β (chr17:46087894-46356512), and γ (chr17:46289349-46707123) were obtained from two previous studies^14,15^ (eFigure 1 in **Supplement**). Segmental duplications can introduce mapping challenges and thus inaccurate calling of the number of copies^21–23^. To address this, we removed segmental duplicated regions inside the α, β and γ (eFigure 5 in **Supplement**) when calculating aligned read depth. Subsequently, the copy numbers of α, β, and γ were obtained based on the aligned read depth on chr17:46135415-46203287, chr17:46106189-46135415, and chr17:46356512-46489410/chr17:46565081-46707123, respectively. Copies of α, β and γ were genotyped by assessing aligned read depth within each 1 Kb bin on the specified regions using CNVpytor (Version 1.3.1)^24^. Then, we employed K-means^25^ to assign an integer copy number for α, β and γ for the 4,076 individuals.

### Validation of CNV calling

To validate the copy numbers of α, β and γ called from WGS, 66 samples were genotyped using TaqMan CNV assay. Overall, the copy number of α, β, or γ inferred by aligned read depth from WGS were highly consistent (α, R = 0.87; γ, R = 0.96) with that from TaqMan assay (eFigure 6 **in Supplement**). The experimental procedure was as follows: Copy number at the α region (Context Sequence: TTAGTCAATTTCTTAGCCAACCCAT; chr17:46171656-46171680, GRCh38) was determined using a predesigned TaqMan CNV assay (Applied Biosystems Part No. Hs01788222_cn). Copy number at the γ region (Context Sequence: AGAAAAAAAGCATTGACTCCAACCC; chr17:46419867-46419891 and chr17:46637445-46637469, GRCh38) was determined using a custom designed TaqMan CNV assay (Applied Biosystems; Forward Primer: TGGCACAATGACCATCGAGATT, Reverse Primer: CTGCCATCTTGTCGGTGTCA, Reporter Sequence: AAGGGTTGGAGTCAATGCTTTT). For the copy number assay reaction, 20 ng of genomic DNA was combined with TaqMan Master Mix (Applied Biosystems Part No. 4371357), 20X TaqMan CNV assay, and 20x RNase P TaqMan Copy Number Reference Assay (Applied Biosystems Part No. 4403328) in a total volume of 20ul. The DNA amplification and quantification were carried out in a QuantStudio 12K Flex instrument (Applied Biosystems) in a 96 well format with the following program: 50°C 2min, 95°C 10min followed by 40 cycles of 95°C 15sec, 60°C 1min. The results were analyzed using the TaqMan CopyCaller software (Version 2.0) (Applied Biosystems).

### Determine haplotypic contributions to diploid copy number

For approximately sixty percent of the samples, only one combination of the structural forms (eFigure 1 in **Supplement**) was possible based on the H1 and H2 genotypes, determined by the 238-bp deletion, and the copy numbers of α, β and γ. For the rest of the samples, multiple haplotypic combinations were possible. Therefore, we initially assigned an equal likelihood to all possible haplotypic combinations that were consistent with the detected copy number of α, β, and γ. Then, the following expectation-maximization (EM) loop described in the previous study^14^ were repeated: from the probabilistic inferences of structural form of 17q21.31 in each sample, we estimated an allele frequency for each structural form of 17q21.31; we then re-estimated the relative likelihood of each combination of haplotypes in each sample, given the population-level allele frequency. In this way, we intended to eliminate haplotypic combinations that are theoretically possible but extremely unlikely given the apparent frequencies of haplotypes as estimated from the rest of the population. The revised probabilistic estimates then allowed a new population-level estimate of haplotype frequencies. We repeated this EM loop process until estimates of haplotype frequency converged. The allele frequency of each structural form of 17q21.31 after convergence were showed in eFigure 1 in **Supplement**.

### Phasing for MAPT sub-haplotypes

The six SNVs (rs1467967, rs242557, rs3785883, rs2471738, rs8070723, and rs7521)^11–13^ on *MAPT* were employed to defined the 26 *MAPT* sub-haplotypes (eTable 2 in **Supplement**). We phased the six SNVs with other SNVs and indels in chr17:43000000-48000000 to determine the *MAPT* sub-haplotypes. The SNV genotypes for the study subjects were called in our previous work^26^. Variants were removed if they were monomorphic, did not pass variant quality score recalibration, had an average read depth ≥ 500, or if all calls have DP<10 & GQ<20. Individual calls with a DP<10 or GQ<20 were set to missing. Then, common variants (MAF > 0.01) with 0.25 < ABHet < 0.75 were phased using SHAPEIT4^27^ (Version 4.2.2).

### Linkage disequilibrium between structural forms of 17q21.31 and MAPT sub-haplotypes

To phase the structural forms of 17q21.31 together with *MAPT* sub-haplotypes, we encoded the copy numbers of α, β and γ as multi-allelic CNVs by a series of surrogate bi-allelic markers with 0/1 alleles^14^ (eTable 3 in **Supplement**). Then, SHAPEIT4^27^ (Version 4.2.2) were used for phasing the copy numbers of α, β and γ together with SNVs/indels. SNVs and indels insides α, β, and γ regions (chr17:46087000-46708000) were not included when phasing. After phasing, we calculated the linkage disequilibrium (LD) between structural forms of 17q21.31 and *MAPT* sub-haplotypes.

### Statistical Analysis

Statistical analyses were performed for the 4,076 individuals (N_PSP_ = 1,684, N_control_ = 2,392). For the association of the copy numbers of α, β, and γ with PSP, logistic regression adjusting for sex and PC1-5 for population substracture was employed. Age was not adjusted in the regression model with following considerations: first, more than half of the PSP cases have age missing (**Table 1**); second, controls (average age, 81) was much older compared to PSP (average age, 63) and well over the mean age-of onset for PSP which is 63 years^28^. Individuals with the H2H2 genotype are imbalanced (5 cases, 138 controls) and with few cases, therefore, statistical analysis for this subgroup was not included.

To evaluate the association of the structural forms of 17q21.31 with PSP, each structural form with allele frequency > 1% is compared with the rest of structural forms using logistic regression model adjusting for sex and PC1-5. To evaluate the association of *MAPT* sub-haplotypes with PSP, each *MAPT* sub-haplotypes with allele frequency > 1% was compared with the rest of sub-haplotypes using logistic regression adjusting for sex and PC1-5.

## Results

### The copy numbers of α, β, and γ and PSP risk

The H1 and H2 haplotypes are the most prominent genetic risk factor for PSP. Individuals with the H2 haplotype showed significantly lower risk of disease (OR, 0.19; 95%CI, 0.16-0.22; *P* = 3.00 × 10^-79^). Structural forms of 17q21.31 can be characterized by three large duplications α, β, and γ^14,15^. However, their specific contributions to the risk of PSP have not been carefully assessed. In this study, we first evaluated whether the copy numbers of α, β, or γ are associated to the risk of PSP independently from H1 and H2 (eFigure 7 in **Supplement**).

We found that each additional copy of γ was associated with 1.08 (95%CI, 1.02-1.15; *P* = 0.014) fold of increased risk of PSP in H1H1 individuals and 1.29 (%95CI, 1.06-1.56; *P* = 0.0096) fold of increased risk of PSP in H1H2 individuals (**Table 2**). The association of the copy number of γ with PSP in H2H2 individuals was not evaluated since there were only five H2H2 PSP samples. On average, γ was associated with 1.10 (95% CI, 1.04-1.17; *P* = 0.0018) fold of increased risk of PSP after adjusting for sex, PC1-5, and allele count (0, 1, or 2) of the H2 haplotype (**Table 2**). Without adjusting for H2, the effect of γ would be neglected (OR, 0.98; 95%CI, 0.93-1.04; *P* = 0.60, **Table. 2**) because the H2 haplotype has increased copies (usually two copies) of γ while displayed significantly lower risk of PSP.

**Table 2.** Association between the copy numbers of α, β, γ and risk of PSP.

| Model: Sex, PC1-5<br>(H1H1 carriers, N=2,958<br>(PSP = 1,511; Control = 1,447) | Model: Sex, PC1-5<br>(H1H2 carriers, N=975)<br>(PSP = 168; Control = 807) |
| --- | --- |

|  | OR (95% CI) | P | OR (95% CI) | P |
| --- | --- | --- | --- | --- |
| $\alpha$ | 0.91 (0.81-1.02) | 0.11 | 0.81 (0.58-1.11) | 0.20 |
| $\beta$ | 0.91 (0.81-1.02) | 0.11 | 0.79 (0.53-1.15) | 0.23 |
| $\gamma$ | 1.08 (1.02-1.15) | 0.014* | 1.29 (1.06-1.56) | 0.0096* |
| **Model: Sex, PC1-5**<br>(N = 4,076)<br>(PSP = 1,684; Control = 2,392) |  |  | **Model: Sex, PC1-5, H2**<br>(N = 4,076)<br>(PSP = 1,684; Control = 2,392) |  |
|  | OR (95% CI) | P | OR (95% CI) | P |
| $\alpha$ | 0.57 (0.52-0.63) | $< 2 \times 10^{-16}$* | 0.90 (0.81-1.00) | 0.061 |
| $\beta$ | 1.14 (1.03-1.27) | 0.011* | 0.90 (0.81-1.01) | 0.064 |
| $\gamma$ | 0.98 (0.93-1.04) | 0.60 | 1.10 (1.04-1.17) | 0.0018* |

\*Represents statistical significance ( $< 0.05$ )

We found no significant association for the copy numbers of α (OR, 0.9; 95%CI 0.81-1.00; *P* = 0.061) and β (OR, 0.9; 95%CI 0.81-1.01; *P* =0.064) with PSP when adjusting the allele count (0, 1, or 2) of H2 (**Table 2**); however, individuals with more copies of α and β showed slightly lower odds ratio for PSP. Similarly, no significance was observed for the copy numbers of α and β when association analysis was stratified by H1H1 or H1H2 genotype. Only under the regression model without adjusting H1 and H2, we observed statistically significant association for the copy numbers of α (*P* < 2 × 10^-16^) and β (*P* = 0.011) with PSP. However, the observed significance mainly arises from their correlation with the H1 and H2 haplotypes, i.e., the increased copies (usually two copies) of α and the absence of β duplication in the H2 haplotype.

### Structural forms of 17q21.31 and PSP risk

We then evaluated whether the structural forms of 17q21.31, characterized by the α, β, and γ, show distinct effect on the risk of PSP. We found that the odds ratio for PSP increases from 1.21 (H1β1γ1, 95%CI 1.10-1.33) to 1.57 (H1β1γ4, 95%CI 1.10-2.26) as the copy number of γ increases from one copy to four copies (**Table 3**). With an additional copy of β, H1β2γ1 (OR, 1.24; 95%CI 1.11-1.38; *P* = 1.87 × 10^-4^) displayed similar risk compared to H1β1γ1 (OR, 1.21; 95%CI 1.10-1.33; *P* = 5.47 × 10^-5^) regarding PSP. This reaffirmed our finding that the copy number of γ was associated with increased risk of PSP independently from H1 and H2, and β was not associated with the risk of PSP (**Table 2**; eFigure 7 **in Supplement**). Besides, all structural forms with H1 background displayed increased risk of PSP and all structural forms with H2 background displayed decreased risk of PSP (**Table 3**).

**Table 3.** Structural forms of 17q21.31 and the risk of PSP.

| Structural forms | Frequency (%) |  | Odds ratio | P |
| --- | --- | --- | --- | --- |
|  | PSP (N = 1,684) | Control (N = 2,392) |  |  |
| H1β1γ1 | 32.57 | 28.20 | 1.21 (1.10-1.33) | $5.47 \times 10^{-5}$ |
| H1β1γ2 | 21.94 | 16.99 | 1.29 (1.16-1.43) | $1.35 \times 10^{-6}$ |
| H1β1γ3 | 14.73 | 10.26 | 1.45 (1.27-1.65) | $3.94 \times 10^{-8}$ |
| H1β1γ4 | 1.93 | 1.28 | 1.57 (1.10-2.26) | $1.35 \times 10^{-2}$ |
| H1β2γ1 | 22.98 | 19.69 | 1.24 (1.11-1.38) | $1.87 \times 10^{-4}$ |
| H2α1γ2 | 0.42 | 1.78 | 0.23 (0.12-0.40) | $5.94 \times 10^{-7}$ |
| H2α2γ2 | 4.78 | 20.09 | 0.19 (0.16-0.23) | $< 2 \times 10^{-16}$ |

Haplotypes in less than 1% of individuals were excluded. Odds ratio and $P$ value were from logistic regression adjusting for PC1-5 and Sex.

### Structural forms of 17q21.31 and MAPT sub-haplotypes

According to the LD structure in *MAPT* gene (∼150 Kb), 26 *MAPT* sub-haplotypes can be determined by six haplotype-tagging SNVs^11–13^ (eTable 2 **in Supplement**). We confirmed the association of H1c (OR 1.79; 95%CI 1.58-2.04; *P* = 1.84 × 10^-19^), H1d (OR 1.52; 95%CI 1.29-1.79; *P* = 3.89 × 10^-7^), and H1o (OR 2.88; 95%CI 2.15-3.89; *P* = 2.77 × 10^-12^) with PSP (**Table 4**). Moreover, H1g (OR 1.46; 95%CI 1.07-1.98; *P* = 0.016), which was significant, and H1h (OR 1.36; 95%CI 1.10-1.69; *P* = 0.0053), which was not significant in the previous study^11^, were both nominal significant in our analysis.

**Table 4.**
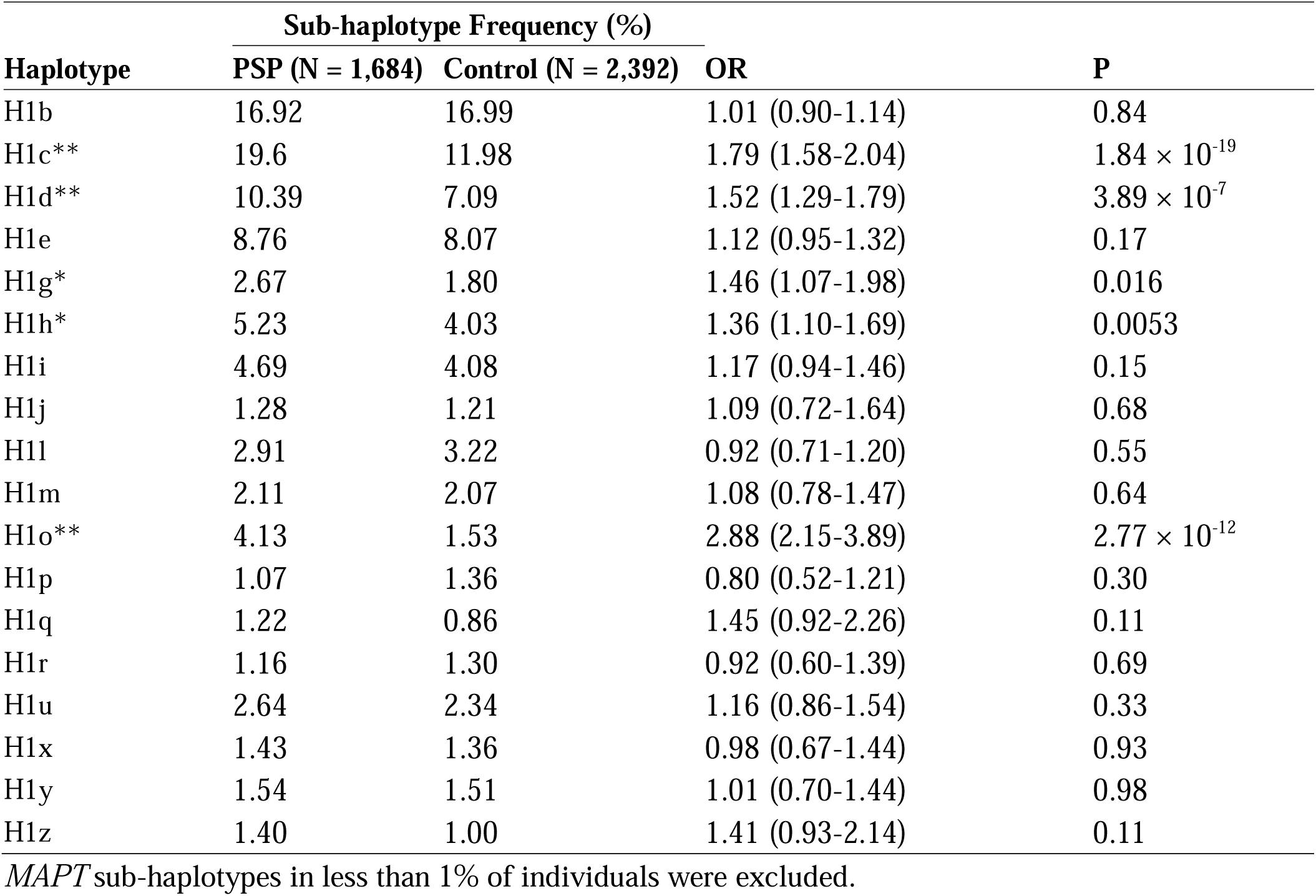
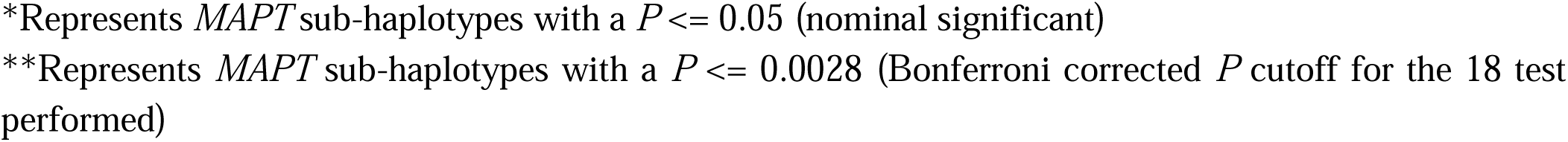
*MAPT* sub-haplotypes defined by six SNVs and the risk of PSP.

| Haplotype | Sub-haplotype Frequency (%) |  | OR | P |
| --- | --- | --- | --- | --- |
|  | PSP (N = 1,684) | Control (N = 2,392) |  |  |
| H1b | 16.92 | 16.99 | 1.01 (0.90-1.14) | 0.84 |
| H1c** | 19.6 | 11.98 | 1.79 (1.58-2.04) | $1.84 \times 10^{-19}$ |
| H1d** | 10.39 | 7.09 | 1.52 (1.29-1.79) | $3.89 \times 10^{-7}$ |
| H1e | 8.76 | 8.07 | 1.12 (0.95-1.32) | 0.17 |
| H1g* | 2.67 | 1.80 | 1.46 (1.07-1.98) | 0.016 |
| H1h* | 5.23 | 4.03 | 1.36 (1.10-1.69) | 0.0053 |
| H1i | 4.69 | 4.08 | 1.17 (0.94-1.46) | 0.15 |
| H1j | 1.28 | 1.21 | 1.09 (0.72-1.64) | 0.68 |
| H1l | 2.91 | 3.22 | 0.92 (0.71-1.20) | 0.55 |
| H1m | 2.11 | 2.07 | 1.08 (0.78-1.47) | 0.64 |
| H1o** | 4.13 | 1.53 | 2.88 (2.15-3.89) | $2.77 \times 10^{-12}$ |
| H1p | 1.07 | 1.36 | 0.80 (0.52-1.21) | 0.30 |
| H1q | 1.22 | 0.86 | 1.45 (0.92-2.26) | 0.11 |
| H1r | 1.16 | 1.30 | 0.92 (0.60-1.39) | 0.69 |
| H1u | 2.64 | 2.34 | 1.16 (0.86-1.54) | 0.33 |
| H1x | 1.43 | 1.36 | 0.98 (0.67-1.44) | 0.93 |
| H1y | 1.54 | 1.51 | 1.01 (0.70-1.44) | 0.98 |
| H1z | 1.40 | 1.00 | 1.41 (0.93-2.14) | 0.11 |

*MAPT* sub-haplotypes in less than 1% of individuals were excluded.
\*Represents *MAPT* sub-haplotypes with a $P \leq 0.05$ (nominal significant) \*\*Represents *MAPT* sub-haplotypes with a $P \leq 0.0028$ (Bonferroni corrected $P$ cutoff for the 18 test performed)

We examined the relationship between the structural forms of 17q21.31 (∼1.5 Mb) and *MAPT* sub-haplotypes (∼150 Kb) through LD analysis. We identified two pairs with R^2^ > 0.1 (eTable 4 in **Supplement**): H1β1γ3 and H1c (R^2^ = 0.31) and H1β2γ1 and H1b (R^2^ = 0.29). This was confirmed by the fact that 70% of H1β1γ3 are H1c and 56% of H1β2γ1 are H1b (**Fig. 1A**). Moreover, with additional copies of γ in the H1, the proportion of *MAPT* sub-haplotypes with increased risk of PSP (i.e., H1c, H1d, H1g, H1h, and H1o) increases from 34% (H1β1γ1) to 77% (H1β1γ4) (**Fig. 1A**). In contrast, the proportion of *MAPT* sub-haplotypes with increased risk of PSP drops from 34% (H1β1γ1) to 11% (H1β2γ1) with an additional copy of β (**Fig. 1A**). In individuals with H1H2 genotypes, no phasing is needed before comparison, therefore, the association between the structural forms of 17q21.31 and *MAPT* sub-haplotypes can be observed directly without phasing (**Fig. 1B**).

**Fig. 1:**
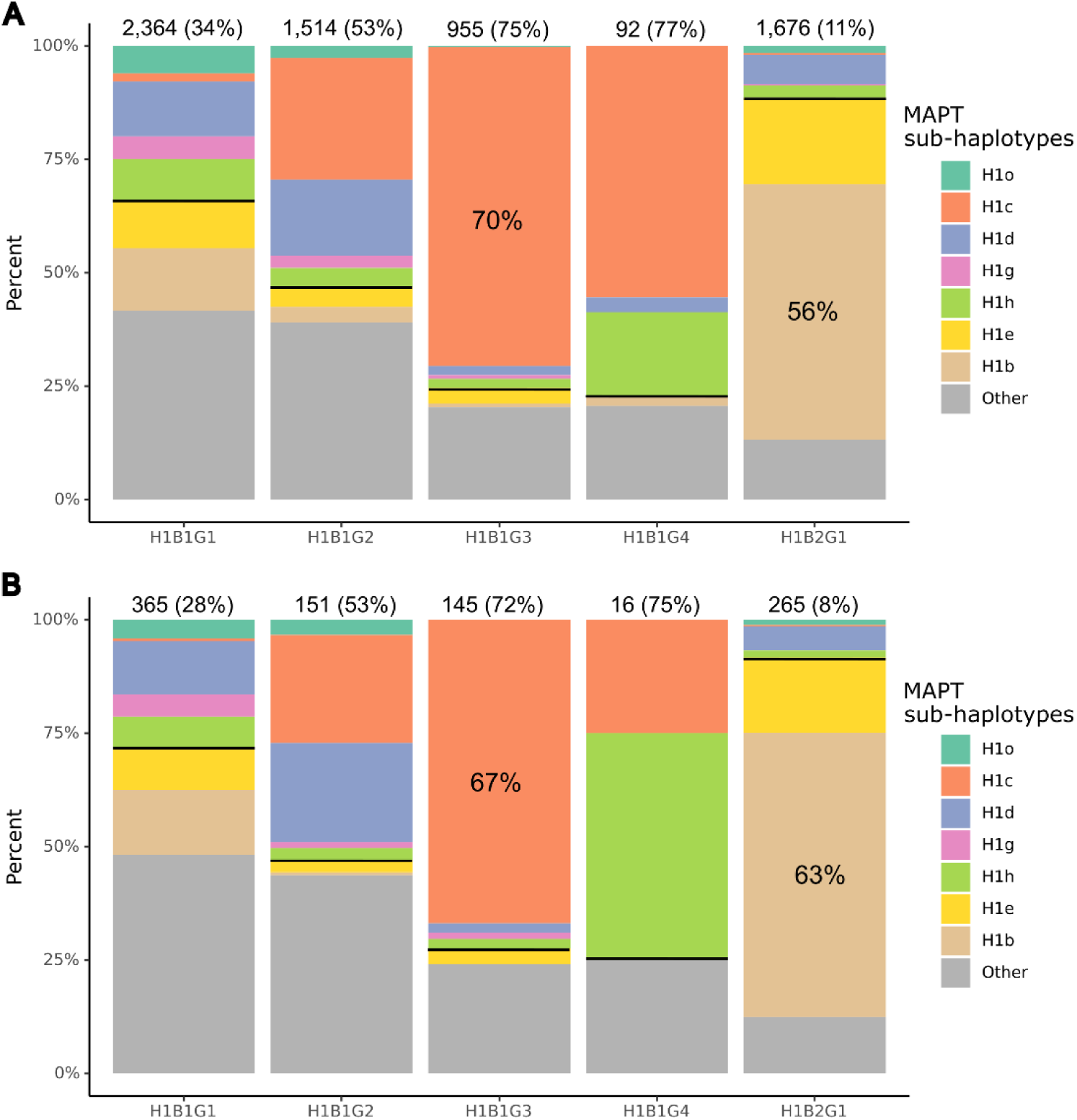
Structural forms of 17q21.31 and *MAPT* sub-haplotypes. Structural forms of 17q.21.31 in less than 1% of individuals were excluded. Structural forms with H2 background are also excluded since there is only one *MAPT* sub-haplotypes for H2. The number above each bar is the total number of *haplotypes* for the specific structural form of 17q21.31 and the percentage that are also *MAPT* sub-haplotypes (H1o, H1c, H1d, H1g, H1h) associated with increased risk of PSP. Besides, the percentage of H1β1γ3 that is H1c and the percentage of of H1β2γ1 that is H1b are displayed. **A.** In phased haplotypes from all samples, the proportion of *MAPT* sub-haplotypes in each structural form of 17q21.31. **B.** In H1H2 individuals, the proportion of *MAPT* sub-haplotypes in each structural form of 17q21.31.

## Discussion

The H1 and H2 haplotypes on 17q21.31 is the strongest genetic risk factor for PSP^7^. The *MAPT* gene inside H1 and H2 showed haplotypic-specific expression and was considered as the possible cause of the association^29,30^. Therefore, previous studies have been focused on variants and haplotypic structures inside *MAPT* and identified sub-haplotypes H1c, H1d, H1o, and H1g, which displayed increased risk of PSP^11,13^. In this study, we went beyond *MAPT* gene (∼150 Kb) and evaluated the association of the structural forms of 17q21.31 (∼1.5 Mb), characterized by large duplications α, β, and γ, with the risk PSP. We found that the copy number of γ duplication was associated with increased risk of PSP and structural forms with γ duplication (such as H1β1γ2, H1β1γ3, and H1β1γ4) had a higher odds ratio for PSP compared to H1β1γ1. This is in accordance with the fact that structural forms with additional copies of γ (from H1β1γ1 to H1β1γ4) tended to have more *MAPT* sub-haplotypes with a significant risk to PSP (i.e., H1c, H1d, H1o, H1g, H1h). Overall, this study provided a first analysis on the association of structural forms of 17q21.31 with the risk of PSP and *MAPT* sub-haplotypes.

It should be noted that the copy numbers of α, β, and γ are correlated with H1 and H2. When performing the association of the copy numbers of α, β, and γ with PSP in individuals with European ancestry, previous studies demonstrated that the 205-Kb β region can only duplicate in the H1 haplotype, the smaller 155-Kb α region but not the entire β region duplicates in the H2 haplotype, and the 210-Kb γ region usually duplicates only once in the H2 haplotype (eFigure 1 in **Supplement**)^15,31^. Therefore, it is important to adjust the status of H1 and H2 and split analysis by individuals with H1H1 and H1H2 genotypes when performing the association of α, β, and γ with PSP. In terms of genomic position, the α region is included in β region, and parts of β and γ are overlapping with each other (eFigure 1 in **Supplement**). When performing the association of α with PSP, we considered whenever there was a β duplication there was also a α duplication as α region is included in β. Considering these factors, we found that only γ is independently associated with risk of PSP, and the associations of α and β with PSP are due to their correlation with H1 and H2.

The influence of structural forms of 17q21.31 on the risk of PSP has never been examined before. In this study, we found that all structural forms with H1 background were association with increased with of PSP. Particularly, H1 with additional γ copies (H1β1γ2, H1β1γ3, and H1β1γ4) displayed a higher odds ratio for PSP compared to H1β1γ1. In previous studies of *MAPT* sub-haplotypes, only a few sub-haplotypes (i.e., H1c, H1d, H1o, and H1g) were associated with increased risk of PSP while other sub-haplotypes displayed no differences against the rest of the population^11,13^. We replicated the association of H1c, H1d, H1o and H1g with PSP and identified H1h with nominal significance. Furthermore, we showed that the H1 haplotype with additional γ copies had more *MAPT* sub-haplotypes that have higher risk of PSP (H1c, H1d, H1o, H1g and H1h), the proportion of *MAPT* sub-haplotypes associated with PSP increased from 34% in H1β1γ1 to 77% in H1β1γ4.

## Limitations

Despite our finding of the importance of the copy number of γ duplication in the risk of PSP, there are a few limitations in the current study. First, all our PSP samples are of European ancestry. It is important to collect samples from other ancestries to validate the discovery in a genetically diverse background. Second, not all PSP cases are pathologically confirmed: of the 1,684 individuals with PSP, 1,386 were autopsy confirmed. Finally, the association analysis needed to be replicated in an independent dataset. Currently, the available resource for whole genome sequencing data in PSP is limited. Therefore, future study focusing on α, β, and γ regions using CNV array to replicate the findings in a large independent cohorts might be helpful.

## Conclusions

In this study of 4076 subjects (N_PSP_ = 1,684, N_control_ = 2,392), we found that γ duplication was associated with increased risk (OR, 1.10; 95% CI, 1.04-1.17; *P* = 0.0018) of PSP. In accordance with this finding, the structural forms with additional copies of γ (H1β1γ2 [OR, 1.29; 95%CI 1.16-1.43; *P* = 1.35 × 10^-6^], H1β1γ3 [OR, 1.45; 95%CI 1.27-1.65; P = 3.94 × 10^-8^], H1β1γ4[OR, 1.57; 95%CI 1.10-2.26; P = 1.35 × 10^-2^]) showed higher odds ratio for PSP compared to H1β1γ1 (OR, 1.21; 95%CI 1.10-1.33; *P* = 5.47 × 10^-5^) and had higher proportion of *MAPT* sub-haplotypes associated with increased risk of PSP (i.e., H1c, H1d, H1g, H1o, and H1g).

## Data Availability

The data is available at https://www.niagads.org/home

## Declarations

### Ethics approval and consent to participate

### Consent for publication

Not applicable.

### Availability of data and materials

NIAGADS Data Sharing Service (https://dss.niagads.org/)

## Acknowledgements

This project is supported by CurePSP, courtesy of a donation from the Morton and Marcine Friedman Foundation. We are indebted to the Biobanc-Hospital Clinic-FRCB-IDIBAPS and Center for Neurodegenerative Disease Research at Penn for samples and data procurement. The PSP genetics study group is a multisite collaboration including: German Center for Neurodegenerative Diseases (DZNE), Munich; Department of Neurology, LMU Hospital, Ludwig-Maximilians-Universität (LMU), Munich, Germany (Franziska Hopfner, Günter Höglinger); German Center for Neurodegenerative Diseases (DZNE), Munich; Center for Neuropathology and Prion Research, LMU Hospital, Ludwig-Maximilians-Universität (LMU), Munich, Germany (Sigrun Roeber, Jochen Herms); Justus-Liebig-Universität Gießen, Germany (Ulrich Müller); MRC Centre for Neurodegeneration Research, King’s College London, London, UK (Claire Troakes); Movement Disorders Unit, Neurology Department and Neurological Tissue Bank and Neurology Department, Hospital Clínic de Barcelona, University of Barcelona, Barcelona, Catalonia, Spain (Ellen Gelpi; Yaroslau Compta); Department of Neurology and Netherlands Brain Bank, Erasmus Medical Centre, Rotterdam, The Netherlands (John C. van Swieten); Division of Neurology, Royal University Hospital, University of Saskatchewan, Canada (Alex Rajput); Australian Brain Bank Network in collaboration with the Victorian Brain Bank Network, Australia (Fairlie Hinton), Department of Neurology, Hospital Ramón y Cajal, Madrid, Spain (Justo García de Yebenes). The acknowledgement of PSP cohorts is listed below, whereas the acknowledgement of ADSP cohorts for control samples can be found in the supplementary materials. The Genotype-Tissue Expression (GTEx) Project was supported by the Common Fund of the Office of the Director of the National Institutes of Health, and by NCI, NHGRI, NHLBI, NIDA, NIMH, and NINDS. The data used for the analyses described in this manuscript were obtained from: https://gtexportal.org/home/datasets the GTEx Portal on 1/27/2022. We also thank to Drs. Murray Grossman and Hans Kretzschmar for their valuable contribution to this work.

<u>AMP-AD (sa000011) data:</u> Mayo RNAseq Study-Study data were provided by the following sources: The Mayo Clinic Alzheimer’s Disease Genetic Studies, led by Dr. Nilufer Ertekin-Taner and Dr. Steven G. Younkin, Mayo Clinic, Jacksonville, FL using samples from the Mayo Clinic Study of Aging, the Mayo Clinic Alzheimer’s Disease Research Center, and the Mayo Clinic Brain Bank. Data collection was supported through funding by NIA grants P50 AG016574, R01 AG032990, U01 AG046139, R01 AG018023, U01 AG006576, U01 AG006786, R01 AG025711, R01 AG017216, R01 AG003949, NINDS grant R01 NS080820, CurePSP Foundation, and support from Mayo Foundation. Study data includes samples collected through the Sun Health Research Institute Brain and Body Donation Program of Sun City, Arizona. The Brain and Body Donation Program is supported by the National Institute of Neurological Disorders and Stroke (U24 NS072026 National Brain and Tissue Resource for Parkinson’s Disease and Related Disorders), the National Institute on Aging (P30 AG19610 Arizona Alzheimer’s Disease Core Center), the Arizona Department of Health Services (contract 211002, Arizona Alzheimer’s Research Center), the Arizona Biomedical Research Commission (contracts 4001, 0011, 05-901 and 1001 to the Arizona Parkinson’s Disease Consortium) and the Michael J. Fox Foundation for Parkinson’s Research.

<u>PSP-NIH-CurePSP-Tau (sa000015) data</u>: This project was funded by the NIH grant UG3NS104095 and supported by grants U54NS100693 and U54AG052427. Queen Square Brain Bank is supported by the Reta Lila Weston Institute for Neurological Studies and the Medical Research Council UK. The Mayo Clinic Florida had support from a Morris K. Udall Parkinson’s Disease Research Center of Excellence (NINDS P50 #NS072187), CurePSP and the Tau Consortium. The samples from the University of Pennsylvania are supported by NIA grant P01AG017586.

<u>PSP-CurePSP-Tau (sa000016) data</u>: This project was funded by the Tau Consortium, Rainwater Charitable Foundation, and CurePSP. It was also supported by NINDS grant U54NS100693 and NIA grants U54NS100693 and U54AG052427. Queen Square Brain Bank is supported by the Reta Lila Weston Institute for Neurological Studies and the Medical Research Council UK. The Mayo Clinic Florida had support from a Morris K. Udall Parkinson’s Disease Research Center of Excellence (NINDS P50 #NS072187), CurePSP and the Tau Consortium. The samples from the University of Pennsylvania are supported by NIA grant P01AG017586. Tissues were received from the Victorian Brain Bank, supported by The Florey Institute of Neuroscience and Mental Health, The Alfred and the Victorian Forensic Institute of Medicine and funded in part by Parkinson’s Victoria and MND Victoria. We are grateful to the Sun Health Research Institute Brain and Body Donation Program of Sun City, Arizona for the provision of human biological materials (or specific description, e.g. brain tissue, cerebrospinal fluid). The Brain and Body Donation Program is supported by the National Institute of Neurological Disorders and Stroke (U24 NS072026 National Brain and Tissue Resource for Parkinson’s Disease and Related Disorders), the National Institute on Aging (P30 AG19610 Arizona Alzheimer’s Disease Core Center), the Arizona Department of Health Services (contract 211002, Arizona Alzheimer’s Research Center), the Arizona Biomedical Research Commission (contracts 4001, 0011, 05-901 and 1001 to the Arizona Parkinson’s Disease Consortium) and the Michael J. Fox Foundation for Parkinson’s Research. Biomaterial was provided by the Study Group DESCRIBE of theClinical Research of the German Center for Neurodegenerative Diseases (DZNE).

PSP_UCLA (sa000017) data: Thank to the AL-108-231 investigators, Adam L Boxer, Anthony E Lang, Murray Grossman, David S Knopman, Bruce L Miller, Lon S Schneider, Rachelle S Doody, Andrew Lees, Lawrence I Golbe, David R Williams, Jean-Cristophe Corvol, Albert Ludolph, David Burn, Stefan Lorenzl, Irene Litvan, Erik D Roberson, Günter U Höglinger, Mary Koestler, Cliff ord R Jack Jr, Viviana Van Deerlin, Christopher Randolph, Iryna V Lobach, Hilary W Heuer, Illana Gozes, Lesley Parker, Steve Whitaker, Joe Hirman, Alistair J Stewart, Michael Gold, and Bruce H Morimoto.

## Authors’ information

### Authors’ contribution

Study design: TSC, DD, GUH, JYT, DHG, GDS, and WPL. Sample collection, brain biospecimens, and neuropathological examinations: TSC, CM, LM, AR, PPDD, NLB, MG, LDK, JCVS, ED, BFG, KLN, CT, JGdY, ARG, TM, WHO, GR, UM, FH, TA, SR, PP, AB, AD, ILB, TGC, GES, LNH, IL, RR, OR, DG, ALB, BLM, WWS, VMVD, EBL, CLW, HM, JH, RdS, JFC, AMG, GC, and DHG. Genotype or phenotype acquisition: HW, TSC, VP, LVB, KF, AN, LSW, DHG, GDS, and WPL. Variant detection and variant quality check: HW, TSC, VP, LVB, KF, YYL, and WPL. Statistical analyses and interpretation of results: HW, YQS, AT, TSC, KF, AN, JYT, DHG, GDS, and WPL. Experimental validation: BAD and PLC. Draft of the manuscript: HW, GDS, and WPL. All authors read, critically revised, and approved the manuscript.

