## Supplementary Information for "Association of Structural Forms of 17q21.31 with the Risk of Progressive Supranuclear Palsy and *MAPT* Sub-haplotypes"

eTable 1. Whole genome sequencing of study subjects

|  | **Overall (N = 4,618)** | **Control (N = 2,821)** | **PSP (N = 1,797)** |
| --- | --- | --- | --- |
| **PCR** |  |  |  |
| PCRFree | 3113 (67.41%) | 2443 (86.60%) | 670 (37.28%) |
| PCRAmplified | 1505 (32.59%) | 378 (13.40%) | 1127 (62.72%) |
| **Sequencing Platform** |  |  |  |
| Illumina_HiSeqX | 3550 (76.87%) | 2007 (71.14%) | 1543 (85.87%) |
| Illumina_HiSeq_2000/2500 | 850 (18.41%) | 596 (21.13%) | 254 (14.13%) |
| Illumina_NovaSeq | 218 (4.72%) | 218 (7.73%) | 0 (0.00%) |
| **Sequencing Center** |  |  |  |
| USUHS | 1895 (41.04%) | 1308 (46.37%) | 587 (32.67%) |
| MACROGEN | 873 (18.90%) | 0 (0.00%) | 873 (48.58%) |
| Illumina | 711 (15.40%) | 522 (18.50%) | 189 (10.52%) |
| NYGC | 620 (13.43%) | 472 (16.73%) | 148 (8.24%) |
| Broad | 361 (7.82%) | 361 (12.80%) | 0 (0.00%) |
| Baylor | 84 (1.82%) | 84 (2.98%) | 0 (0.00%) |
| WashU | 62 (1.34%) | 62 (2.20%) | 0 (0.00%) |
| GENENTECH | 12 (0.26%) | 12 (0.43%) | 0 (0.00%) |

eTable 2. MAPT sub-haplotypes defined by six SNVs

| **Haplotype** | **rs1467967** | **rs242557** | **rs3785883** | **rs2471738** | **rs8070723** | **rs7521** |
| --- | --- | --- | --- | --- | --- | --- |
| H1b | G | G | G | C | A | A |
| H1c | A | A | G | T | A | G |
| H1d | A | A | G | C | A | A |
| H1e | A | G | G | C | A | A |
| H1f | G | G | A | C | A | A |
| H1g | G | A | A | C | A | A |
| H1h | A | G | A | C | A | A |
| H1i | G | A | G | C | A | A |
| H1j | A | G | G | C | A | G |
| H1k | A | A | A | C | A | G |
| H1l | A | G | A | C | A | G |
| H1m | G | A | G | C | A | G |
| H1n | G | G | A | C | A | G |
| H1o | A | A | A | C | A | A |
| H1p | G | G | G | T | A | G |
| H1q | A | A | G | T | A | A |
| H1r | A | G | G | T | A | G |
| H1s | G | G | G | C | A | G |
| H1t | A | G | A | T | A | G |
| H1u | A | A | G | C | A | G |
| H1v | G | G | A | T | A | G |
| H2w | G | G | G | C | G | G |
| H1x | G | A | A | T | A | G |
| H1y | A | A | A | T | A | G |
| H1z | G | A | G | T | A | G |
| H2 | A | G | G | C | G | G |

eTable 3. Surrogate biallelic encoding of the structural forms of 17q21.31

| **Haplotype** | **H1/H2** | **R1H1 (β)** | **R2H2 (α)** | **R3H1 (γ on H1)** | **R3H2 (γ on H2)** |
| --- | --- | --- | --- | --- | --- |
| H1β1γ1 | 0 | 100 | 00 | 1000 | 00 |
| H1β1γ2 | 0 | 100 | 00 | 1100 | 00 |
| H1β1γ3 | 0 | 100 | 00 | 1110 | 00 |
| H1β1γ4 | 0 | 100 | 00 | 1111 | 00 |
| H1β2γ1 | 0 | 110 | 00 | 1000 | 00 |
| H1β3γ1 | 0 | 111 | 00 | 1000 | 00 |
| H2α1γ1 | 1 | 000 | 10 | 0000 | 10 |
| H2α1γ2 | 1 | 000 | 10 | 0000 | 11 |
| H2α2γ1 | 1 | 000 | 11 | 0000 | 10 |
| H2α2γ2 | 1 | 000 | 11 | 0000 | 11 |

eTable 4. Linkage disequilibrium between structural forms of 17q21.31 and MAPT sub-haplotypes

| **Haplotype (SV)** | **Haplotype (SNV)** | **D** | **D_prime** | **R2** |
| --- | --- | --- | --- | --- |
| H1β1γ3 | H1c | 0.065982661 | 0.638079472 | 0.313110902 |
| H1β2γ1 | H1b | 0.082503498 | 0.607260883 | 0.289179089 |
| H1β1γ1 | H2 | -0.046467149 | 0.998482112 | 0.078355742 |
| H1β1γ1 | H1c | -0.040384828 | 0.883269341 | 0.060047106 |
| H1β2γ1 | H2 | -0.032539217 | 0.997730025 | 0.048635771 |
| H1β2γ1 | H1c | -0.030940024 | 0.965622217 | 0.044612826 |
| H1β1γ2 | H2 | -0.029676361 | 0.997625335 | 0.043344367 |
| H1β2γ1 | H1e | 0.023320629 | 0.34859269 | 0.042452147 |
| H1β1γ2 | H1c | 0.025943249 | 0.20931738 | 0.033607554 |
| H1β1γ4 | H1c | 0.008031492 | 0.610779815 | 0.032531966 |
| H1β1γ1 | H1l | 0.014065899 | 0.640532154 | 0.031157543 |
| H1β1γ2 | H1b | -0.024888347 | 0.75923184 | 0.028195773 |
| H1β1γ3 | H2 | -0.018991083 | 0.997571985 | 0.025565906 |
| H1β1γ2 | H1y | 0.007671692 | 0.615479319 | 0.025133391 |
| H1β1γ3 | H1b | -0.019335351 | 0.921654435 | 0.024510174 |
| H1β1γ1 | H1i | 0.014471762 | 0.470457814 | 0.023853857 |
| H1β2γ1 | H1q | 0.005869226 | 0.728604919 | 0.020652531 |
| H1β1γ1 | H1o | 0.010121529 | 0.547878481 | 0.019079329 |
| H1β1γ1 | H1h | 0.012940211 | 0.402428713 | 0.01828313 |
| H1β1γ2 | H1v | 0.004765826 | 0.64069375 | 0.016151174 |
| H1β1γ1 | H1g | 0.008044939 | 0.524546586 | 0.014452714 |
| H1β1γ2 | H1d | 0.012939987 | 0.186835162 | 0.013856584 |
| H1β1γ1 | H1d | 0.013637647 | 0.227140305 | 0.011348405 |
| H1β1γ3 | H1p | 0.003807372 | 0.345669139 | 0.010914697 |
| H1β1γ2 | H1r | 0.004386138 | 0.43202102 | 0.010057156 |
| H1β1γ1 | H1m | 0.006426899 | 0.433836941 | 0.009542008 |
| H1β1γ1 | H1n | 0.003854728 | 0.680541072 | 0.008859539 |
| H1β1γ2 | H1x | 0.004177905 | 0.36781049 | 0.008168209 |
| H1β1γ2 | H1p | 0.003875231 | 0.381698264 | 0.007850653 |
| H1β1γ3 | H1z | 0.002985196 | 0.288141599 | 0.007128151 |
| H1β1γ1 | H1k | 0.002321135 | 0.783421472 | 0.006117531 |
| H1β1γ3 | H1d | -0.006891412 | 0.659367076 | 0.005660507 |
| H1β1γ3 | H1i | -0.004998181 | 0.933416601 | 0.005558053 |
| H1β2γ1 | H1l | -0.005089219 | 0.777141742 | 0.005162894 |
| H1β1γ3 | H1r | 0.002383175 | 0.216367082 | 0.004276346 |
| H1β2γ1 | H1g | -0.003820636 | 0.835358122 | 0.004126084 |
| H1β1γ1 | H1p | -0.003252426 | 0.868408519 | 0.004077491 |
| H1β1γ1 | H1r | -0.00323997 | 0.865082659 | 0.004046319 |
| H1β1γ1 | H1t | 0.002166521 | 0.540479365 | 0.003945236 |
| H1β1γ1 | H1f | 0.002696457 | 0.423881578 | 0.003863993 |
| H1β1γ1 | H1j | 0.003059444 | 0.347612253 | 0.003607971 |
| H1β1γ3 | H1l | -0.0033701 | 0.881617966 | 0.003493788 |
| H1β2γ1 | H1p | -0.002563342 | 0.976640792 | 0.00320593 |
| H1β2γ1 | H1j | -0.002563342 | 0.976640792 | 0.00320593 |
| H1β1γ2 | H1e | -0.006152775 | 0.381175072 | 0.003166146 |
| H1β2γ1 | H1m | -0.003273788 | 0.741056545 | 0.003134013 |
| H1β1γ4 | H1b | -0.002589693 | 0.970751092 | 0.003083317 |
| H1β1γ3 | H1o | -0.002910108 | 0.904922677 | 0.00308086 |
| H1β2γ1 | H1y | -0.002732514 | 0.847989193 | 0.002975937 |
| H1β1γ4 | H2 | -0.002342789 | 0.967767936 | 0.002728396 |
| H1β1γ3 | H1h | -0.003547108 | 0.633703806 | 0.002683488 |
| H1β1γ3 | H1e | -0.004643766 | 0.449532841 | 0.002597645 |
| H1β2γ1 | H1z | -0.002176143 | 0.881482204 | 0.00245463 |
| H1β1γ4 | H1h | 0.001261972 | 0.085175292 | 0.002381932 |
| H1β1γ2 | H1j | 0.001990062 | 0.196014953 | 0.00207035 |
| H1β1γ1 | H1x | 0.002388051 | 0.242515303 | 0.001967725 |
| H1β1γ4 | H1p | 0.00060542 | 0.049014865 | 0.001935324 |
| H1β2γ1 | H1x | -0.002076059 | 0.706986262 | 0.001882433 |
| H1β2γ1 | H1u | 0.002747087 | 0.139123826 | 0.00187374 |
| H1β2γ1 | H1h | -0.003665793 | 0.382288561 | 0.001857234 |
| H1β1γ3 | H1x | -0.001643579 | 0.958849196 | 0.001820711 |
| H1β2γ1 | H1v | -0.001565243 | 0.813953943 | 0.001626004 |
| H1β2γ1 | H1r | -0.001822286 | 0.694296452 | 0.001620221 |
| H1β2γ1 | H1d | -0.00456085 | 0.254727738 | 0.00160661 |
| H1β1γ1 | H1u | 0.002746179 | 0.156785798 | 0.00147931 |
| H1β2γ1 | H1n | -0.001378806 | 0.816280575 | 0.001434805 |
| H1β1γ1 | H1e | 0.004821981 | 0.081255427 | 0.001433862 |
| H1β1γ3 | H1q | -0.001201754 | 0.966139456 | 0.001336173 |
| H1β2γ1 | H1s | -0.001324961 | 0.772518266 | 0.001305018 |
| H1β1γ4 | H1e | -0.001235579 | 0.940600235 | 0.001289613 |
| H1β1γ1 | H1s | 0.0014262 | 0.247976502 | 0.001194561 |
| H1β2γ1 | H1i | -0.002818323 | 0.307231493 | 0.001145144 |
| H1β1γ2 | H1s | 0.001181366 | 0.178067403 | 0.001111601 |
| H1β1γ3 | H1f | -0.001033618 | 0.933416601 | 0.001109054 |
| H1β1γ4 | H2w | 4.55E-05 | 0.372217841 | 0.001091294 |
| H1β1γ2 | H1q | -0.001282769 | 0.659986553 | 0.001057008 |
| H1β1γ3 | H1y | -0.001307037 | 0.694871624 | 0.00105074 |
| H1β1γ3 | H1g | -0.001549634 | 0.580436583 | 0.001047479 |
| H1β2γ1 | H1k | -0.000834343 | 0.944312164 | 0.001000525 |
| H1β1γ3 | H1n | -0.000920473 | 0.93354697 | 0.0009868 |
| H1β2γ1 | H1t | -0.000957874 | 0.801309031 | 0.000976172 |
| H1β2γ1 | H1o | -0.002004257 | 0.363803858 | 0.00094698 |
| H1β1γ1 | H1b | -0.005269157 | 0.102744106 | 0.00093184 |
| H1β1γ4 | H1z | 0.000378643 | 0.032591075 | 0.000804211 |
| H1β1γ4 | H1d | -0.000946636 | 0.712271526 | 0.000749004 |
| H1β1γ3 | H1t | -0.000622607 | 0.892265798 | 0.00063644 |
| H1β1γ4 | H1i | -0.000602887 | 0.885407253 | 0.000567088 |
| H1β1γ3 | H1k | -0.00046655 | 0.904602398 | 0.000482786 |
| H1β1γ4 | H1m | 0.000362849 | 0.023871621 | 0.000416631 |
| H1β1γ4 | H1l | -0.000408064 | 0.839479207 | 0.00035921 |
| H1β1γ2 | H1g | 0.001030249 | 0.058233574 | 0.000321455 |
| H1β1γ2 | H1z | 0.000710841 | 0.074437649 | 0.000280624 |
| H1β1γ4 | H1o | -0.000330907 | 0.809192265 | 0.000279348 |
| H1β2γ1 | H1f | -0.00063468 | 0.334565679 | 0.00027097 |
| H1β1γ4 | H1g | -0.000261465 | 0.77016341 | 0.000209119 |
| H1β1γ3 | H1m | -0.000634699 | 0.246125614 | 0.000181784 |
| H1β1γ2 | H1f | 0.000451139 | 0.061479684 | 0.000146691 |
| H1β1γ3 | H1u | -0.000611506 | 0.20055934 | 0.00014328 |
| H1β1γ1 | H1q | -0.000537556 | 0.176785926 | 0.000136866 |
| H1β1γ2 | H1l | -0.000732959 | 0.122709866 | 0.000114741 |
| H1β1γ4 | H1y | -0.000161785 | 0.676390081 | 0.000112895 |
| H1β1γ3 | H1v | -0.000317634 | 0.282965311 | 0.000103331 |
| H1β1γ4 | H1x | -0.000147433 | 0.676390081 | 0.000102737 |
| H1β1γ4 | H1q | -0.000116056 | 0.733730724 | 8.74E-05 |
| H1β1γ3 | H2w | 3.23E-05 | 0.295999206 | 7.83E-05 |
| H1β1γ4 | H1r | 0.000112951 | 0.009144559 | 6.74E-05 |
| H1β1γ4 | H1j | 0.000112951 | 0.009144559 | 6.74E-05 |
| H1β1γ2 | H1h | 0.000632875 | 0.017062223 | 5.93E-05 |
| H1β1γ2 | H1o | 0.000478898 | 0.022472493 | 5.79E-05 |
| H1β1γ2 | H1u | -0.000448715 | 0.094183581 | 5.36E-05 |
| H1β1γ1 | H1v | -0.000307254 | 0.111970371 | 4.95E-05 |
| H1β1γ1 | H1y | -0.000381242 | 0.082911835 | 4.58E-05 |
| H1β1γ4 | H1v | -6.80E-05 | 0.476388152 | 3.32E-05 |
| H1β1γ4 | H1f | -6.71E-05 | 0.476388152 | 3.28E-05 |
| H1β1γ4 | H1s | -6.06E-05 | 0.476388152 | 2.96E-05 |
| H1β1γ2 | H2w | 2.37E-05 | 0.236233711 | 2.94E-05 |
| H1β2γ1 | H2w | 2.15E-05 | 0.218477284 | 2.24E-05 |
| H1β1γ2 | H1n | -0.000161217 | 0.104640357 | 2.10E-05 |
| H1β1γ3 | H1s | -0.000119625 | 0.119486279 | 1.64E-05 |
| H1β1γ1 | H1z | -0.000157797 | 0.044793349 | 1.02E-05 |
| H1β1γ2 | H1i | 0.000221323 | 0.006237289 | 7.57E-06 |
| H1β1γ2 | H1t | 6.87E-05 | 0.014855547 | 5.38E-06 |
| H1β1γ4 | H1k | -1.64E-05 | 0.249817473 | 4.18E-06 |
| H1β1γ1 | H2w | 1.04E-05 | 0.118970496 | 4.13E-06 |
| H1β1γ2 | H1m | 0.000111561 | 0.006528437 | 3.90E-06 |
| H1β1γ3 | H1j | -6.98E-05 | 0.045585354 | 3.67E-06 |
| H1β1γ4 | H1t | -1.36E-05 | 0.152778233 | 2.12E-06 |
| H1β1γ4 | H1u | -1.65E-05 | 0.042506565 | 7.30E-07 |
| H1β1γ4 | H1n | 5.79E-06 | 0.000728483 | 2.74E-07 |
| H1β1γ2 | H1k | -1.18E-05 | 0.014656189 | 2.15E-07 |

eFigure 1. Structural forms of 17q21.31 haplotypes defined by the copy number of α, β, and γ.

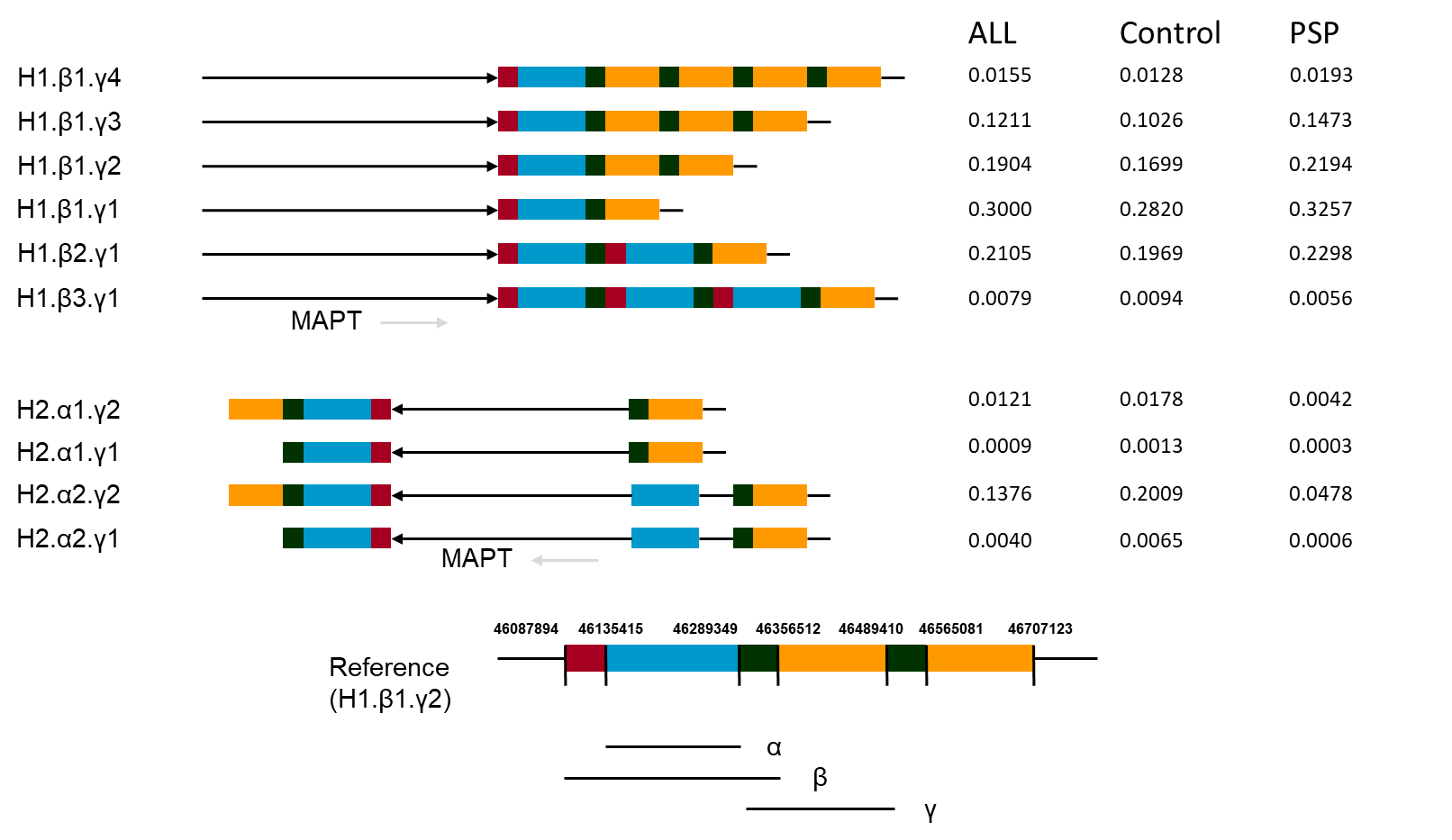

The 205-Kb β region can only be duplicated in H1 haplotypes, the smaller 155-Kb α region but not the entire β region is duplicated in H2 haplotypes, and the 210-Kb γ region usually duplicate only once in H2 haplotypes.

eFigure 2. The aligned read depth on α, β，and γ region

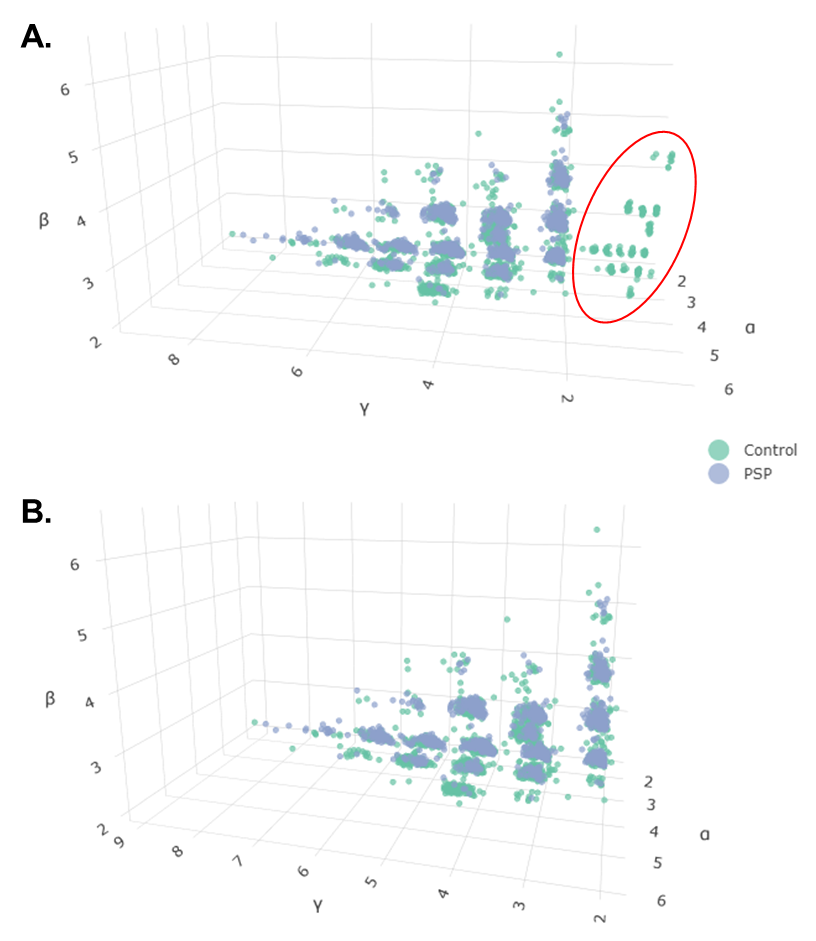

**A.** The read depth (obtained by CNVpytor) on α, β, and γ region. The red eclipse shows that there are 183 samples with abnormally low coverage (aligned read depth < 1.7) on γ region. **B.** The coverage on α, β, and γ region after removing outliers.

eFigure 3. The copy number of α, β，and γ by H1/H2 haplotypes.

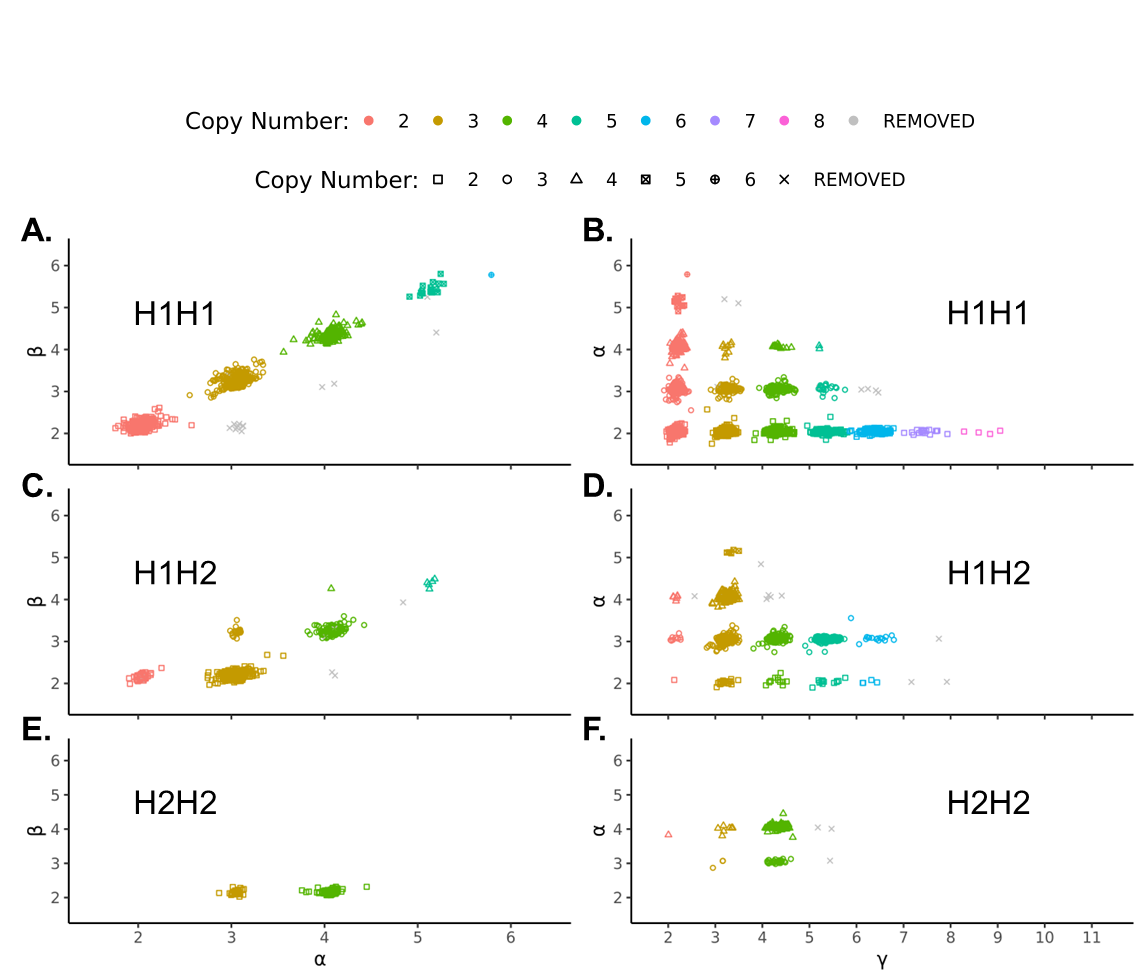

**A.** Copy number of α against β in individuals with two copies of H1. **B.** Copy number of γ against α in individuals with two copies of H1. **C.** Copy number of α against β in individuals with one H1 haplotype and one H2 haplotype. **D.** Copy number of γ against α in individuals with one H1 haplotype and one H2 haplotype. **E.** Copy number of α against β in individuals with two copies of H2. **F.** Copy number of γ against α in individuals with two copies of H2. “REMOVED” represents individuals removed due to the copy number of α, β, or γ is not possible giving the H1/H2 specification and existing structural forms of H1/H2.

eFigure 4. The copy number of α, β，and γ by H1/H2 haplotypes.

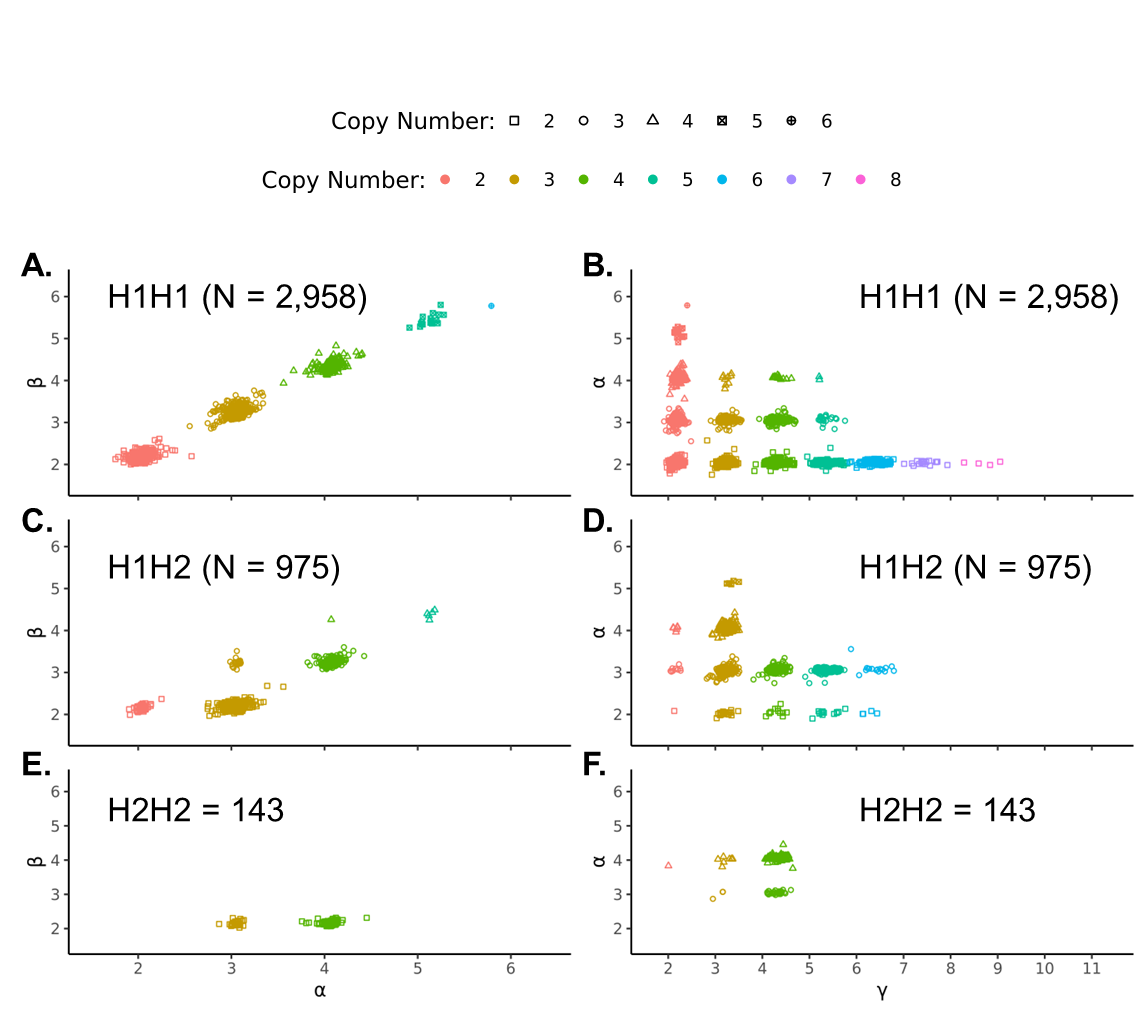

**A.** Copy number of α against β in individuals with two copies of H1. **B.** Copy number of γ against α in individuals with two copies of H1. **C.** Copy number of α against β in individuals with one H1 haplotype and one H2 haplotype. **D.** Copy number of γ against α in individuals with one H1 haplotype and one H2 haplotype. **E.** Copy number of α against β in individuals with two copies of H2. **F.** Copy number of γ against α in individuals with two copies of H2. We excluded 30 individuals (from eFigure 3) whose copy number of α, β and γ were not possible giving the genotype of H1/H2 (by the 238-bp deletion) for that individual and the existing structural form of H1/H2.

eFigure 5. Segmental duplications inside α, β, and γ region

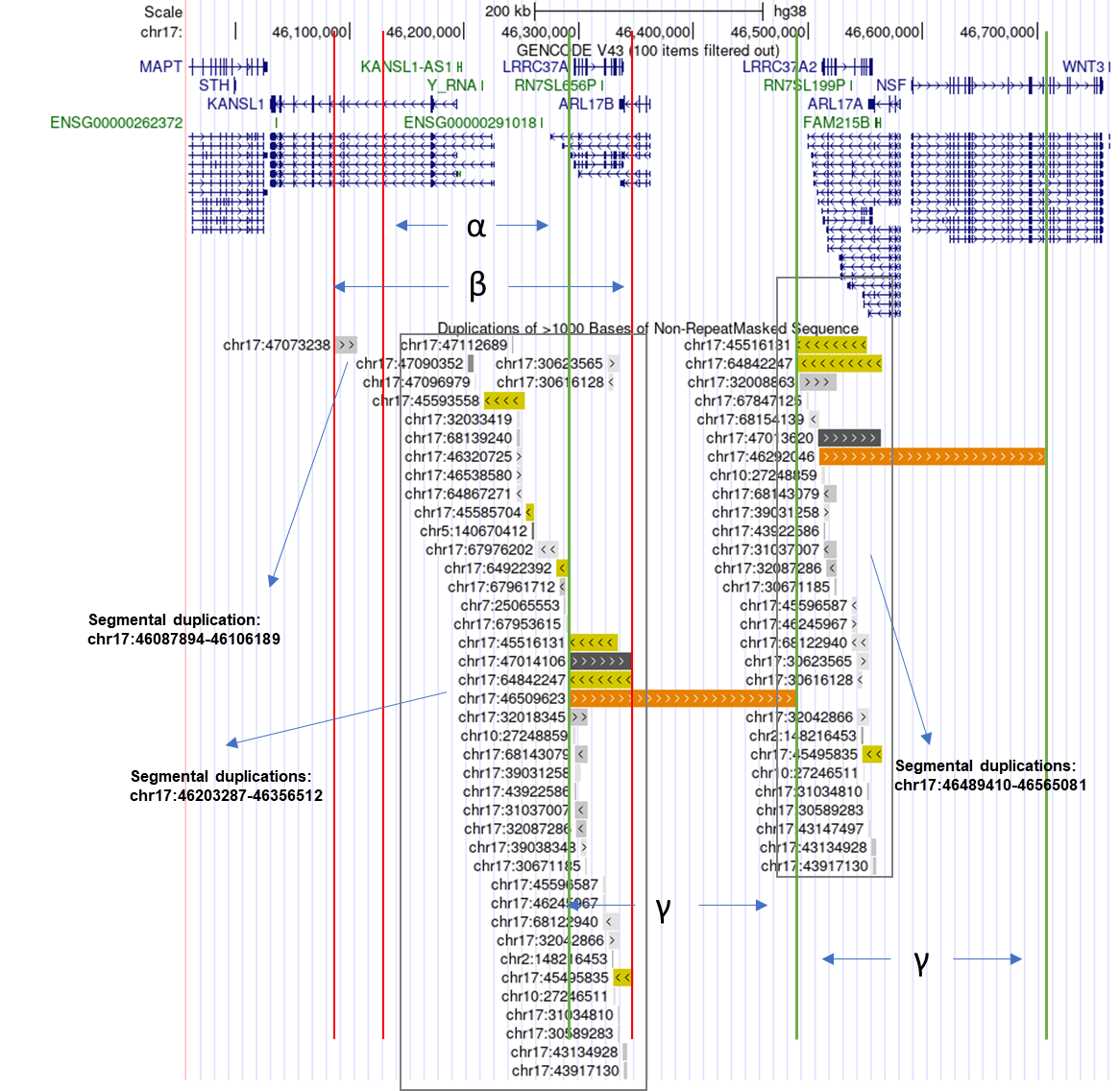

eFigure 6. The copy number of α and γ from WGS and from TaqMan CNV assay

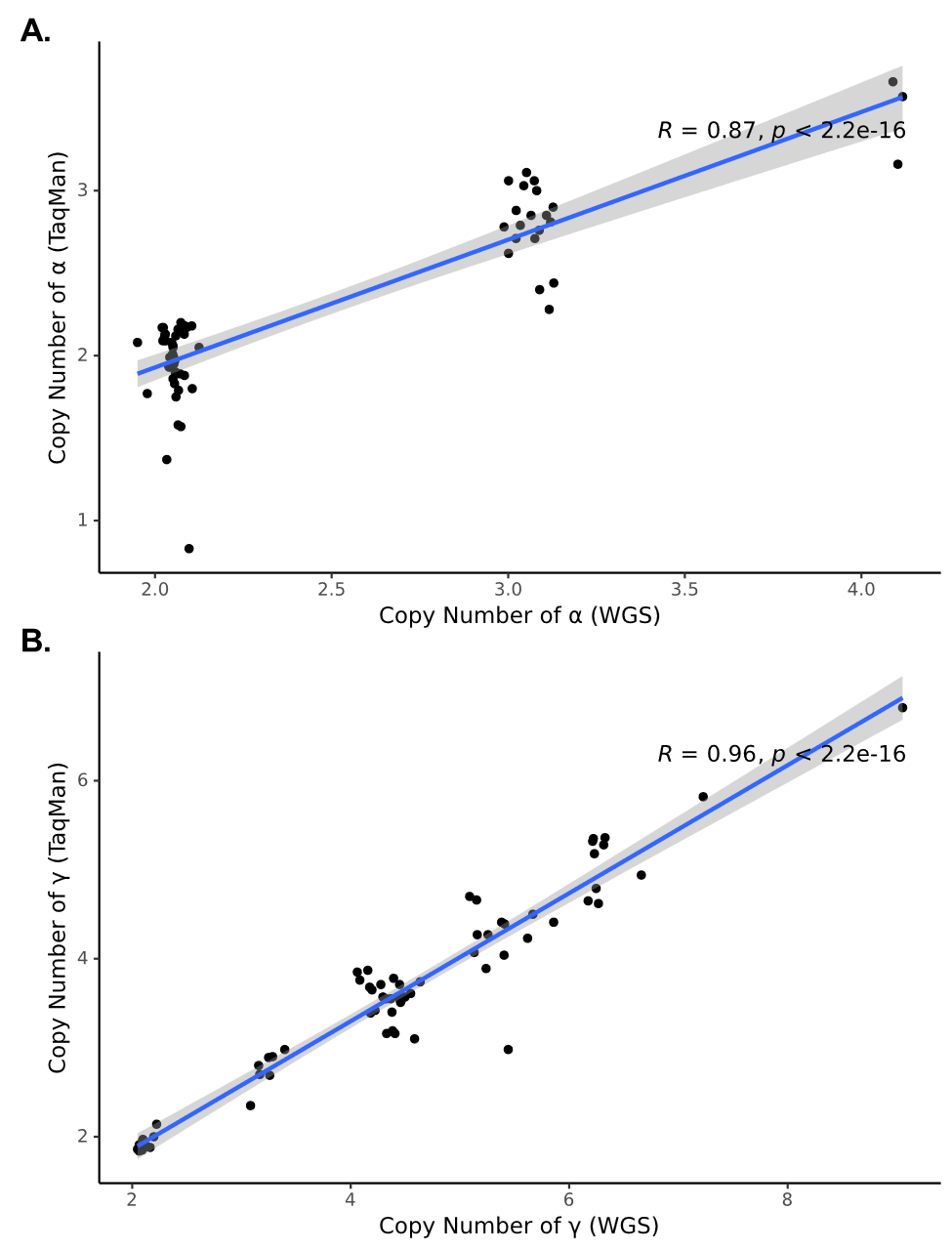

**A.** The copy number of α from WGS and from TaqMan CNV assay. **B.** The copy number of γ from WGS and from TaqMan CNV assay.

eFigure 7: The copy number of α, β，and γ in PSPs and Controls.

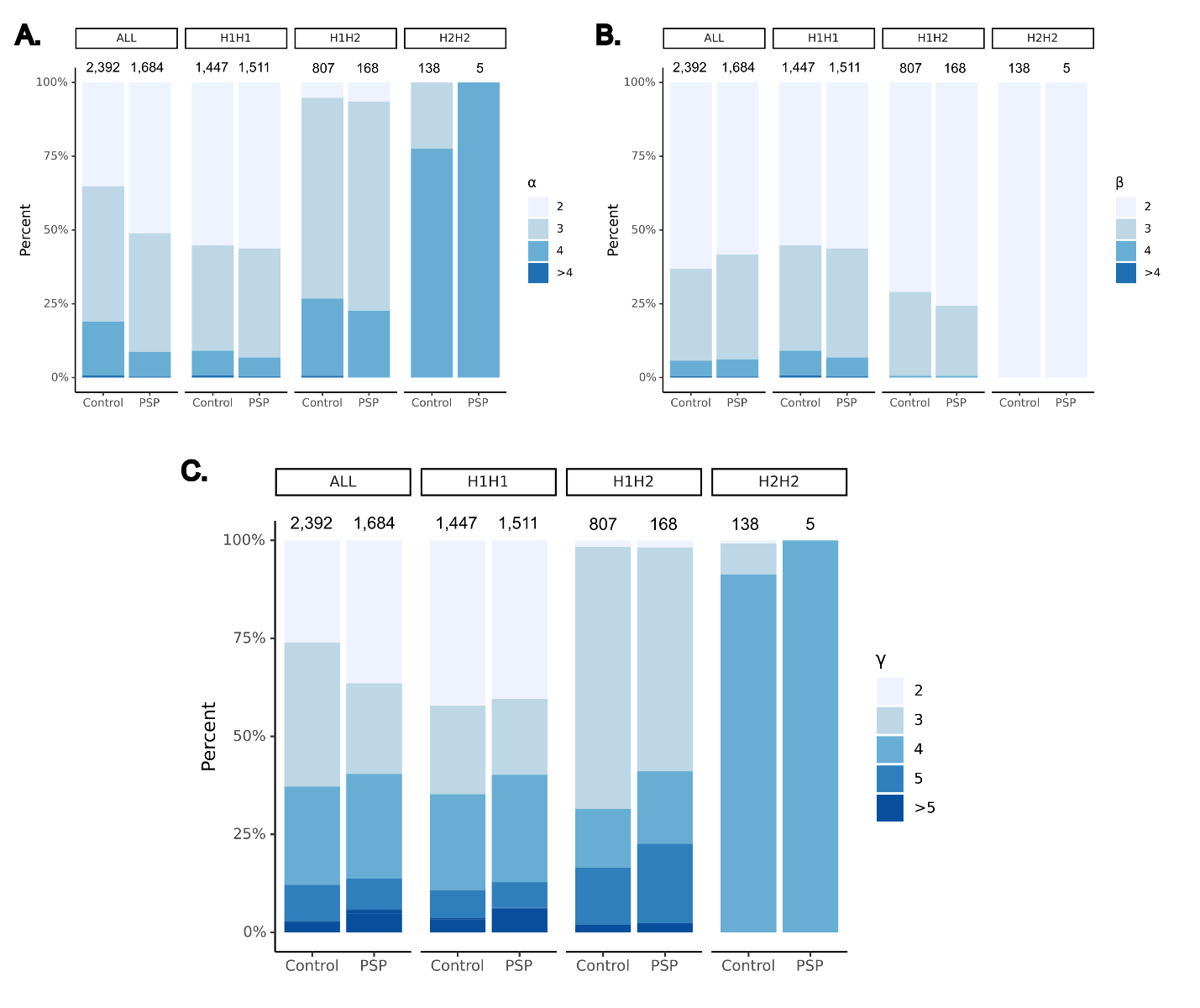

**A.** The copy number of α in all cases/controls and cases/controls stratified by H1H1, H1H2, and H2H2. **B.** The copy number of β in all cases/controls and cases/controls stratified by H1H1, H1H2, and H2H2. **C.** The copy number of γ in all cases/controls and cases/controls stratified by H1H1, H1H2, and H2H2.

Acknowledgements

### ADSP (sa000001) data:

The Alzheimer’s Disease Sequencing Project (ADSP) is comprised of two Alzheimer’s Disease (AD) genetics consortia and three National Human Genome Research Institute (NHGRI) funded Large Scale Sequencing and Analysis Centers (LSAC). The two AD genetics consortia are the Alzheimer’s Disease Genetics Consortium (ADGC) funded by NIA (U01 AG032984), and the Cohorts for Heart and Aging Research in Genomic Epidemiology (CHARGE) funded by NIA (R01 AG033193), the National Heart, Lung, and Blood Institute (NHLBI), other National Institute of Health (NIH) institutes and other foreign governmental and non-governmental organizations. The Discovery Phase analysis of sequence data is supported through UF1AG047133 (to Drs. Schellenberg, Farrer, Pericak-Vance, Mayeux, and Haines); U01AG049505 to Dr. Seshadri; U01AG049506 to Dr. Boerwinkle; U01AG049507 to Dr. Wijsman; and U01AG049508 to Dr. Goate and the Discovery Extension Phase analysis is supported through U01AG052411 to Dr. Goate, U01AG052410 to Dr. Pericak-Vance and U01 AG052409 to Drs. Seshadri and Fornage.

Sequencing for the Follow Up Study (FUS) is supported through U01AG057659 (to Drs. PericakVance, Mayeux, and Vardarajan) and U01AG062943 (to Drs. Pericak-Vance and Mayeux). Data generation and harmonization in the Follow-up Phase is supported by U54AG052427 (to Drs. Schellenberg and Wang). The FUS Phase analysis of sequence data is supported through U01AG058589 (to Drs. Destefano, Boerwinkle, De Jager, Fornage, Seshadri, and Wijsman), U01AG058654 (to Drs. Haines, Bush, Farrer, Martin, and Pericak-Vance), U01AG058635 (to Dr. Goate), RF1AG058066 (to Drs. Haines, Pericak-Vance, and Scott), RF1AG057519 (to Drs. Farrer and Jun), R01AG048927 (to Dr. Farrer), and RF1AG054074 (to Drs. Pericak-Vance and Beecham).

The ADGC cohorts include: Adult Changes in Thought (ACT) (U01 AG006781, U19 AG066567), the Alzheimer’s Disease Research Centers (ADRC) (P30 AG062429, P30 AG066468, P30 AG062421, P30 AG066509, P30 AG066514, P30 AG066530, P30 AG066507, P30 AG066444, P30 AG066518, P30 AG066512, P30 AG066462, P30 AG072979, P30 AG072972, P30 AG072976, P30 AG072975, P30 AG072978, P30 AG072977, P30 AG066519, P30 AG062677, P30 AG079280, P30 AG062422, P30 AG066511, P30 AG072946, P30 AG062715, P30 AG072973, P30 AG066506, P30 AG066508, P30 AG066515, P30 AG072947, P30 AG072931, P30 AG066546, P20 AG068024, P20 AG068053, P20 AG068077, P20 AG068082, P30 AG072958, P30 AG072959), the Chicago Health and Aging Project (CHAP) (R01 AG11101, RC4 AG039085, K23 AG030944), Indiana Memory and Aging Study (IMAS) (R01 AG019771), Indianapolis Ibadan (R01 AG009956, P30 AG010133), the Memory and Aging Project (MAP) ( R01 AG17917), Mayo Clinic (MAYO) (R01 AG032990, U01 AG046139, R01 NS080820, RF1 AG051504, P50 AG016574), Mayo Parkinson’s Disease controls (NS039764, NS071674, 5RC2HG005605), University of Miami (R01 AG027944, R01 AG028786, R01 AG019085, IIRG09133827, A2011048), the Multi-Institutional Research in Alzheimer’s Genetic Epidemiology Study (MIRAGE) (R01 AG09029, R01 AG025259), the National Centralized Repository for Alzheimer’s Disease and Related Dementias (NCRAD) (U24 AG021886), the National Institute on Aging Late Onset Alzheimer’s Disease Family Study (NIA- LOAD) (U24 AG056270), the Religious Orders Study (ROS) (P30 AG10161, R01 AG15819), the Texas Alzheimer’s Research and Care Consortium (TARCC) (funded by the Darrell K Royal Texas Alzheimer’s Initiative), Vanderbilt University/Case Western Reserve University (VAN/CWRU) (R01 AG019757, R01 AG021547, R01 AG027944, R01 AG028786, P01 NS026630, and Alzheimer’s Association), the Washington Heights-Inwood Columbia Aging Project (WHICAP) (RF1 AG054023), the University of Washington Families (VA Research Merit Grant, NIA: P50AG005136, R01AG041797, NINDS: R01NS069719), the Columbia University Hispanic Estudio Familiar de Influencia Genetica de Alzheimer (EFIGA) (RF1 AG015473), the University of Toronto (UT) (funded by Wellcome Trust, Medical Research Council, Canadian Institutes of Health Research), and Genetic Differences (GD) (R01 AG007584). The CHARGE cohorts are supported in part by National Heart, Lung, and Blood Institute (NHLBI) infrastructure grant HL105756 (Psaty), RC2HL102419 (Boerwinkle) and the neurology working group is supported by the National Institute on Aging (NIA) R01 grant AG033193.

The CHARGE cohorts participating in the ADSP include the following: Austrian Stroke Prevention Study (ASPS), ASPS-Family study, and the Prospective Dementia Registry-Austria (ASPS/PRODEM-Aus), the Atherosclerosis Risk in Communities (ARIC) Study, the Cardiovascular Health Study (CHS), the Erasmus Rucphen Family Study (ERF), the Framingham Heart Study (FHS), and the Rotterdam Study (RS). ASPS is funded by the Austrian Science Fond (FWF) grant number P20545-P05 and P13180 and the Medical University of Graz. The ASPS-Fam is funded by the Austrian Science Fund (FWF) project I904), the EU Joint Programme – Neurodegenerative Disease Research (JPND) in frame of the BRIDGET project (Austria, Ministry of Science) and the Medical University of Graz and the Steiermärkische Krankenanstalten Gesellschaft. PRODEM-Austria is supported by the Austrian Research Promotion agency (FFG) (Project No. 827462) and by the Austrian National Bank (Anniversary Fund, project 15435. ARIC research is carried out as a collaborative study supported by NHLBI contracts (HHSN268201100005C, HHSN268201100006C, HHSN268201100007C, HHSN268201100008C, HHSN268201100009C, HHSN268201100010C, HHSN268201100011C, and HHSN268201100012C). Neurocognitive data in ARIC is collected by U01 2U01HL096812, 2U01HL096814, 2U01HL096899, 2U01HL096902, 2U01HL096917 from the NIH (NHLBI, NINDS, NIA and NIDCD), and with previous brain MRI examinations funded by R01-HL70825 from the NHLBI. CHS research was supported by contracts HHSN268201200036C, HHSN268200800007C, N01HC55222, N01HC85079, N01HC85080, N01HC85081, N01HC85082, N01HC85083, N01HC85086, and grants U01HL080295 and U01HL130114 from the NHLBI with additional contribution from the National Institute of Neurological Disorders and Stroke (NINDS). Additional support was provided by R01AG023629, R01AG15928, and R01AG20098 from the NIA. FHS research is supported by NHLBI contracts N01-HC-25195 and HHSN268201500001I. This study was also supported by additional grants from the NIA (R01s AG054076, AG049607 and AG033040 and NINDS (R01 NS017950). The ERF study as a part of EUROSPAN (European Special Populations Research Network) was supported by European Commission FP6 STRP grant number 018947 (LSHG-CT-2006-01947) and also received funding from the European Community’s Seventh Framework Programme (FP7/2007-2013)/grant agreement HEALTH-F4- 2007-201413 by the European Commission under the programme “Quality of Life and Management of the Living Resources” of 5th Framework Programme (no. QLG2-CT-2002- 01254). High-throughput analysis of the ERF data was supported by a joint grant from the Netherlands Organization for Scientific Research and the Russian Foundation for Basic Research (NWO-RFBR 047.017.043). The Rotterdam Study is funded by Erasmus Medical Center and Erasmus University, Rotterdam, the Netherlands Organization for Health Research and Development (ZonMw), the Research Institute for Diseases in the Elderly (RIDE), the Ministry of Education, Culture and Science, the Ministry for Health, Welfare and Sports, the European Commission (DG XII), and the municipality of Rotterdam. Genetic data sets are also supported by the Netherlands Organization of Scientific Research NWO Investments (175.010.2005.011, 911-03-012), the Genetic Laboratory of the Department of Internal Medicine, Erasmus MC, the Research Institute for Diseases in the Elderly (014-93-015; RIDE2), and the Netherlands Genomics Initiative (NGI)/Netherlands Organization for Scientific Research (NWO) Netherlands Consortium for Healthy Aging (NCHA), project 050-060-810. All studies are grateful to their participants, faculty and staff. The content of these manuscripts is solely the responsibility of the authors and does not necessarily represent the official views of the National Institutes of Health or the U.S. Department of Health and Human Services.

The FUS cohorts include: the Alzheimer’s Disease Research Centers (ADRC) (P30 AG062429, P30 AG066468, P30 AG062421, P30 AG066509, P30 AG066514, P30 AG066530, P30 AG066507, P30 AG066444, P30 AG066518, P30 AG066512, P30 AG066462, P30 AG072979, P30 AG072972, P30 AG072976, P30 AG072975, P30 AG072978, P30 AG072977, P30 AG066519, P30 AG062677, P30 AG079280, P30 AG062422, P30 AG066511, P30 AG072946, P30 AG062715, P30 AG072973, P30 AG066506, P30 AG066508, P30 AG066515, P30 AG072947, P30 AG072931, P30 AG066546, P20 AG068024, P20 AG068053, P20 AG068077, P20 AG068082, P30 AG072958, P30 AG072959), Alzheimer’s Disease Neuroimaging Initiative (ADNI) (U19AG024904), Amish Protective Variant Study (RF1AG058066), Cache County Study (R01AG11380, R01AG031272, R01AG21136, RF1AG054052), Case Western Reserve University Brain Bank (CWRUBB) (P50AG008012), Case Western Reserve University Rapid Decline (CWRURD) (RF1AG058267, NU38CK000480), CubanAmerican Alzheimer’s Disease Initiative (CuAADI) (3U01AG052410), Estudio Familiar de Influencia Genetica en Alzheimer (EFIGA) (5R37AG015473, RF1AG015473, R56AG051876), Genetic and Environmental Risk Factors for Alzheimer Disease Among African Americans Study (GenerAAtions) (2R01AG09029, R01AG025259, 2R01AG048927), Gwangju Alzheimer and Related Dementias Study (GARD) (U01AG062602), Hillblom Aging Network (2014-A-004-NET, R01AG032289, R01AG048234), Hussman Institute for Human Genomics Brain Bank (HIHGBB) (R01AG027944, Alzheimer’s Association “Identification of Rare Variants in Alzheimer Disease”), Ibadan Study of Aging (IBADAN) (5R01AG009956), Longevity Genes Project (LGP) and LonGenity (R01AG042188, R01AG044829, R01AG046949, R01AG057909, R01AG061155, P30AG038072), Mexican Health and Aging Study (MHAS) (R01AG018016), Multi-Institutional Research in Alzheimer’s Genetic Epidemiology (MIRAGE) (2R01AG09029, R01AG025259, 2R01AG048927), Northern Manhattan Study (NOMAS) (R01NS29993), Peru Alzheimer’s Disease Initiative (PeADI) (RF1AG054074), Puerto Rican 1066 (PR1066) (Wellcome Trust (GR066133/GR080002), European Research Council (340755)), Puerto Rican Alzheimer Disease Initiative (PRADI) (RF1AG054074), Reasons for Geographic and Racial Differences in Stroke (REGARDS) (U01NS041588), Research in African American Alzheimer Disease Initiative (REAAADI) (U01AG052410), the Religious Orders Study (ROS) (P30 AG10161, P30 AG72975, R01 AG15819, R01 AG42210), the RUSH Memory and Aging Project (MAP) (R01 AG017917, R01 AG42210Stanford Extreme Phenotypes in AD (R01AG060747), University of Miami Brain Endowment Bank (MBB), University of Miami/Case Western/North Carolina A&T African American (UM/CASE/NCAT) (U01AG052410, R01AG028786), and Wisconsin Registry for Alzheimer’s Prevention (WRAP) (R01AG027161 and R01AG054047).

The four LSACs are: the Human Genome Sequencing Center at the Baylor College of Medicine (U54 HG003273), the Broad Institute Genome Center (U54HG003067), The American Genome Center at the Uniformed Services University of the Health Sciences (U01AG057659), and the Washington University Genome Institute (U54HG003079). Genotyping and sequencing for the ADSP FUS is also conducted at John P. Hussman Institute for Human Genomics (HIHG) Center for Genome Technology (CGT).

Biological samples and associated phenotypic data used in primary data analyses were stored at Study Investigators institutions, and at the National Centralized Repository for Alzheimer’s Disease and Related Dementias (NCRAD, U24AG021886) at Indiana University funded by NIA. Associated Phenotypic Data used in primary and secondary data analyses were provided by Study Investigators, the NIA funded Alzheimer’s Disease Centers (ADCs), and the National Alzheimer’s Coordinating Center (NACC, U24AG072122) and the National Institute on Aging Genetics of Alzheimer’s Disease Data Storage Site (NIAGADS, U24AG041689) at the University of Pennsylvania, funded by NIA. Harmonized phenotypes were provided by the ADSP Phenotype Harmonization Consortium (ADSP-PHC), funded by NIA (U24 AG074855, U01 AG068057 and R01 AG059716) and Ultrascale Machine Learning to Empower Discovery in Alzheimer’s Disease Biobanks (AI4AD, U01 AG068057). This research was supported in part by the Intramural Research Program of the National Institutes of health, National Library of Medicine. Contributors to the Genetic Analysis Data included Study Investigators on projects that were individually funded by NIA, and other NIH institutes, and by private U.S. organizations, or foreign governmental or nongovernmental organizations.

### ADNI (sa000002) data:

Data collection and sharing for this project was funded by the Alzheimer's Disease Neuroimaging Initiative (ADNI) (National Institutes of Health Grant U01 AG024904) and DOD ADNI (Department of Defense award number W81XWH-12-2-0012). ADNI is funded by the National Institute on Aging, the National Institute of Biomedical Imaging and Bioengineering, and through generous contributions from the following: AbbVie, Alzheimer’s Association; Alzheimer’s Drug Discovery Foundation; Araclon Biotech; BioClinica, Inc.; Biogen; Bristol-Myers Squibb Company; CereSpir, Inc.; Cogstate; Eisai Inc.; Elan Pharmaceuticals, Inc.; Eli Lilly and Company; EuroImmun; F. Hoffmann-La Roche Ltd and its affiliated company Genentech, Inc.; Fujirebio; GE Healthcare; IXICO Ltd.; Janssen Alzheimer Immunotherapy Research & Development, LLC.; Johnson & Johnson Pharmaceutical Research & Development LLC.; Lumosity; Lundbeck; Merck & Co., Inc.; Meso Scale Diagnostics, LLC.; NeuroRx Research; Neurotrack Technologies; Novartis Pharmaceuticals Corporation; Pfizer Inc.; Piramal Imaging; Servier; Takeda Pharmaceutical Company; and Transition Therapeutics. The Canadian Institutes of Health Research is providing funds to support ADNI clinical sites in Canada. Private sector contributions are facilitated by the Foundation for the National Institutes of Health (www.fnih.org). The grantee organization is the Northern California Institute for Research and Education, and the study is coordinated by the Alzheimer’s Therapeutic Research Institute at the University of Southern California. ADNI data are disseminated by the Laboratory for Neuro Imaging at the University of Southern California.

Additional information to include in an acknowledgment statement can be found on the LONI site: https://adni.loni.usc.edu/wp-content/uploads/how_to_apply/ADNI_Data_Use_Agreement.pdf.

### FASe_Families (sa000004) data:

This work was supported by grants from the National Institutes of Health (R01AG044546, P01AG003991, RF1AG053303, R01AG058501, U01AG058922, RF1AG058501 and R01AG057777). The recruitment and clinical characterization of research participants at Washington University were supported by NIH P50 AG05681, P01 AG03991, and P01 AG026276. This work was supported by access to equipment made possible by the Hope Center for Neurological Disorders, and the Departments of Neurology and Psychiatry at Washington University School of Medicine.

We thank the contributors who collected samples used in this study, as well as patients and their families, whose help and participation made this work possible. This work was supported by access to equipment made possible by the Hope Center for Neurological Disorders, and the Departments of Neurology and Psychiatry at Washington University School of Medicine

### KnightADRC (sa000008) data:

This work was supported by grants from the National Institutes of Health (R01AG044546, P01AG003991, RF1AG053303, R01AG058501, U01AG058922, RF1AG058501 and R01AG057777). The recruitment and clinical characterization of research participants at Washington University were supported by NIH P50 AG05681, P01 AG03991, and P01 AG026276. This work was supported by access to equipment made possible by the Hope Center for Neurological Disorders, and the Departments of Neurology and Psychiatry at Washington University School of Medicine.

We thank the contributors who collected samples used in this study, as well as patients and their families, whose help and participation made this work possible. This work was supported by access to equipment made possible by the Hope Center for Neurological Disorders, and the Departments of Neurology and Psychiatry at Washington University School of Medicine.

### AMP-AD (sa000011) data:

Mayo RNAseq Study- Study data were provided by the following sources: The Mayo Clinic Alzheimer's Disease Genetic Studies, led by Dr. Nilufer Ertekin-Taner and Dr. Steven G. Younkin, Mayo Clinic, Jacksonville, FL using samples from the Mayo Clinic Study of Aging, the Mayo Clinic Alzheimer's Disease Research Center, and the Mayo Clinic Brain Bank. Data collection was supported through funding by NIA grants P50 AG016574, R01 AG032990, U01 AG046139, R01 AG018023, U01 AG006576, U01 AG006786, R01 AG025711, R01 AG017216, R01 AG003949, NINDS grant R01 NS080820, CurePSP Foundation, and support from Mayo Foundation. Study data includes samples collected through the Sun Health Research Institute Brain and Body Donation Program of Sun City, Arizona. The Brain and Body Donation Program is supported by the National Institute of Neurological Disorders and Stroke (U24 NS072026 National Brain and Tissue Resource for Parkinson's Disease and Related Disorders), the National Institute on Aging (P30 AG19610 Arizona Alzheimer's Disease Core Center), the Arizona Department of Health Services (contract 211002, Arizona Alzheimer's Research Center), the Arizona Biomedical Research Commission (contracts 4001, 0011, 05-901 and 1001 to the Arizona Parkinson's Disease Consortium) and the Michael J. Fox Foundation for Parkinson's Research

ROSMAP- We are grateful to the participants in the Religious Order Study, the Memory and Aging Project. This work is supported by the US National Institutes of Health [U01 AG046152, R01 AG043617, R01 AG042210, R01 AG036042, R01 AG036836, R01 AG032990, R01 AG18023, RC2 AG036547, P50 AG016574, U01 ES017155, KL2 RR024151, K25 AG041906-01, R01 AG30146, P30 AG10161, R01 AG17917, R01 AG15819, K08 AG034290, P30 AG10161 and R01 AG11101.

Mount Sinai Brain Bank (MSBB)- This work was supported by the grants R01AG046170, RF1AG054014, RF1AG057440 and R01AG057907 from the NIH/National Institute on Aging (NIA). R01AG046170 is a component of the AMP-AD Target Discovery and Preclinical Validation Project. Brain tissue collection and characterization was supported by NIH HHSN271201300031C.

### UPitt Kamboh (sa000012) data:

This study was supported by the National Institute on Aging (NIA) grants AG030653, AG041718, AG064877 and P30-AG066468.

### NACC Genentech (sa000013) data:

We would like to thank study participants, their families, and the sample collectors for their invaluable contributions. This research was supported in part by the National Institute on Aging grant U01AG049508 (PI Alison M. Goate). This research was supported in part by Genentech, Inc. (PI Alison M. Goate, Robert R. Graham).

The NACC database is funded by NIA/NIH Grant U01 AG016976. NACC data are contributed by these NIA-funded ADCs: P30 AG013846 (PI Neil Kowall, MD), P50 AG008702 (PI Scott Small, MD), P50 AG025688 (PI Allan Levey, MD, PhD), P30 AG010133 (PI Andrew Saykin, PsyD), P50 AG005146 (PI Marilyn Albert, PhD), P50 AG005134 (PI Bradley Hyman, MD, PhD), P50 AG016574 (PI Ronald Petersen, MD, PhD), P30 AG013854 (PI M. Marsel Mesulam, MD), P30 AG008017 (PI Jeffrey Kaye, MD), P30 AG010161 (PI David Bennett, MD), P30 AG010129 (PI Charles DeCarli, MD), P50 AG016573 (PI Frank LaFerla, PhD), P50 AG005131 (PI Douglas Galasko, MD), P30 AG028383 (PI Linda Van Eldik, PhD), P30 AG010124 (PI John Trojanowski, MD, PhD), P50 AG005142 (PI Helena Chui, MD), P30 AG012300 (PI Roger Rosenberg, MD), P50 AG005136 (PI Thomas Grabowski, MD), P50 AG005681 (PI John Morris, MD), P30 AG028377 (Kathleen Welsh-Bohmer, PhD), and P50 AG008671 (PI Henry Paulson, MD, PhD).

Samples from the National Cell Repository for Alzheimer’s Disease (NCRAD), which receives government support under a cooperative agreement grant (U24 AG21886) awarded by the National Institute on Aging (NIA), were used in this study. We thank contributors who collected samples used in this study, as well as patients and their families, whose help and participation made this work possible.

The Alzheimer's Disease Genetics Consortium supported the collection of samples used in this study through National Institute on Aging (NIA) grants U01AG032984 and RC2AG036528.

### CacheCounty (sa000014) data:

We acknowledge the generous contributions of the Cache County Memory Study participants. Sequencing for this study was funded by RF1AG054052 (PI: John S.K. Kauwe).
